# Clusters of multiple long-term conditions in three UK datasets: a latent class analysis

**DOI:** 10.1101/2023.09.05.23294158

**Authors:** Lewis Steell, Stefanie J. Krauth, Sayem Ahmed, Grace Dibben, Emma McIntosh, Peter Hanlon, Jim Lewsey, Barbara I. Nicholl, David McAllister, Rod S. Taylor, Sally J. Singh, Frances S. Mair, Bhautesh D. Jani

## Abstract

**Introduction:** Latent class analysis (LCA) can be used to identify subgroups within populations based on unobserved variables. LCA can be used to explore whether certain long-term conditions (LTC) occur together more frequently than others in patients with multiple-long term conditions. In this manuscript we present findings from applying LCA in three large-scale UK databanks.

**Methods:** We applied LCA to three different UK databanks: Secure Anonymised Information Linkage databank [SAIL], UK Biobank, and Understanding Society: the UK Household Longitudinal Study [UKHLS] and four different age groups: 18-36, 37-54, 55-73, and 74+ years. The optimal number of classes in each LCA was determined using maximum likelihood. Sample size adjusted Bayesian Information Criterion (aBIC) was used to assess model fit and elbow plots and model entropy were used to assess the best number of latent classes in each model.

**Results:** Between three to six clusters were identified in the different datasets and age groups. Although different in detail, similar types of clusters were identified between datasets and age groups which combine disorders around similar systems incl. Cardiometabolic clusters, Pulmonary clusters, Mental health clusters, Painful conditions clusters, and cancer clusters.

## Introduction

Latent class analysis (LCA) is a method of finite mixture modelling used to identify unobserved subgroups (i.e. ‘latent classes’) within a population, based on a series of observed indicator variables [1]. LCA assumes that the distribution of the indicator variables is due to the existence of a finite number of underlying latent classes in the population. Typically applied in behavioural and social sciences, LCA has gained traction in medicine as a means of clustering patients into underlying disease phenotypes, based on their clinical and demographic characteristics. LCA has also been used as a method for identifying clusters of multiple long-term conditions that exist across different populations [2–6]. Identifying clusters of multiple LTCs may facilitate deeper understanding and extraction of the underlying cause and effects of multimorbidity profiles on patient and healthcare outcomes.

LCA facilitates the use of statistical inference in the selection of the optimal model solution and does not rely on arbitrary distance-based measures used in other methods for cluster identification (e.g. k-means or hierarchical clustering). As such, LCA is regarded as a more statistically robust and reproducible method of clustering. Additionally, in simulation studies where the underlying distribution of latent classes was known but concealed during analyses, LCA performed better than other clustering algorithms at accurately classifying individuals into the correct cluster [4, 7]. In clustering at the individual rather than condition level, LCA may uncover a more accurate reflection of the clinical landscape whereby highly prevalent conditions can appear across several identified multiple long-term condition clusters.

Therefore, in this analysis we apply LCA to identify and characterise clusters of multiple long-term conditions across three UK population datasets.

## Methods

### Data source

This study is part of an ongoing NIHR-funded Research project “Personalised exercise rehabilitation for people with multiple long-term conditions (PERFORM)”. The UK Biobank has full ethical approval from the NHS National Research Ethics Service (16/NW/0274). This study was conducted as part of UK Biobank Project 14151. The use and analysis of SAIL data was approved by the SAIL information governance review panel (Project 0830). UKHLS data access and use was granted by the UK Data Service (Project ID: 221571).

This work uses data provided by patients and collected by the NHS as part of their care and support, copyright © (2022), NHS England. Re-used with the permission of the NHS England and UK Biobank. All rights reserved.

This research used data assets made available by National Safe Haven as part of the Data and Connectivity National Core Study, led by Health Data Research UK in partnership with the Office for National Statistics and funded by UK Research and Innovation (research which commenced between 1st October 2020 – 31st March 2021 grant ref MC_PC_20029; 1st April 2021 -30th September 2022 grant ref MC_PC_20058)

### Approach

We applied LCA to three UK datasets (Secure Anonymised Information Linkage databank [SAIL], UK Biobank, and Understanding Society: the UK Household Longitudinal Study [UKHLS]) to identify clusters of multiple long-term conditions in people with multimorbidity. To account for different morbidity profiles across the lifespan, LCA was applied separately to the following age strata: 18 – 36 years, 37 – 54 years, 55 – 73 years, and 74+ years. LCA analyses were applied participants aged 18+ years at baseline data in both research datasets (UK Biobank & UKHLS), and to adults (aged 18+ years) registered with a participating practice on 1^st^ January 2011 in the unselected community cohort (SAIL). This date was chosen as *de facto* ‘baseline’ in SAIL as electronic data capture was most complete after this period, plus it coincided approximately with baseline recruitment of the other datasets [8].

For LCA, input indicators were single LTCs that were derived from either self-report (UK Biobank & UKHLS) or from Read codes [with associated prescription data for some LTCs] from individually linked healthcare data (SAIL). In our analyses, LTCs were deemed present if self-reported or recorded in linked health data, or otherwise absent, meaning there were no missing input data in our LCA models. Due to heterogeneity of data collection across these datasets, different LTCs were used in the definition of multimorbidity and subsequent construction of LCA models. In UK Biobank and SAIL, multimorbidity was defined using 43 LTCs derived from a list commonly applied in multimorbidity research [9] (Table 1). In UKHLS, data were collected on only 17 LTCs, some of which were merged to map with LTCs on the longer list (e.g. ‘hyperthyroidism’ and ‘hypothyroidism’ were merged to become ‘thyroid disease’). Resultantly, a total of 13 LTCs were used to define multimorbidity in UKHLS (Table 2). Only individuals with 2 or more LTCs (i.e. multimorbidity) were included in LCA. The number of people included in LCA per dataset and for each age group is presented in Table 1.

**Table 1.** Number of participants with multimorbidity included in LCA models per age group in each datase.

| Age Strata | Datasets |  |  |
| --- | --- | --- | --- |
|  | SAIL (n) | UK Biobank (n) | UKHLS (n) |
| 18 - 36 years | 53,818 | - | 689 |
| 37 – 54 years | 139,943 | 42,149 | 2,184 |
| 55 – 73 years | 246,893 | 123,008 | 4,117 |
| 74+ years | 152,905 | - | 1,886 |
| Total | 593,559 | 165,157 | 8,876 |

UK Biobank included in only 2 of 4 age strata due to age limits of participants. n reflects number of people with multimorbidity (i.e. 2 or more LTCs)

### LCA Parameters

LCA was conducted in R using the *poLCA* package [10]. LCA was applied iteratively to identify the optimal number of latent classes in each age group and dataset combination (n = 10). For each combination, we constructed and compared LCA models containing 1 to 10 latent classes. As described above, our indicator variables were single LTCs recorded in each of the datasets (n=43 SAIL & UK Biobank; n=13 UKHLS). *poLCA* uses the expectation-maximization (EM) algorithm with a Newton-Raphson step to identify the maximum likelihood estimates of the model parameters. In our analyses, the EM algorithm was set to complete a maximum of 5000 iterations to achieve model convergence on the maximum likelihood solution. Each LCA model was repeated 25 times to improve the search for a global, rather than local, maximum solution. Model parameters were then validated against models with increasing number of random start values (n = 50 & 100). If the maximum likelihood solution (i.e. the single largest log-likelihood value) could not be replicated from increasing repetitions of model fitting, this indicated poor fit of the data to the specified model and the model was rejected.

### Model selection criteria & cluster assignment

Identifying the optimal latent class solution requires careful consideration of multiple criteria. Here, we interpreted the best model through a combined assessment of model fit data with substantive knowledge and clinical interpretability of clusters. Firstly, we used the Bayesian Information Criteria (BIC) and sample size adjusted BIC (aBIC) to guide model selection. These criteria are derived from the maximum likelihood solution and award model parsimony, with lower values indicating better model fit. Simulation studies have suggested the BIC to be the most reliable model fit indicator, particularly in large samples [11]. Despite this, in LCA models with large sample sizes and high number of indicators (as in our SAIL & UK Biobank analyses), the BIC tends to favour more complex models with a higher number of classes. The BIC was therefore not definitive in our model selection and used as a guide only. We plotted the model fit data using elbow plots, to identify where any subsequent increases in number of latent classes resulted in comparatively lesser improvement in BIC, as has been suggested previously [12]. In addition to this, we assessed the underlying distribution of LTCs within the latent classes for clinical interpretability. Where models may have had similar performance in terms of statistical criteria, the substantive clinical interpretation and face validity of identified clusters were considered prior to model selection. Finally, we considered model entropy, with higher values (close to 1) representing models with more accurate assignment of individuals into the appropriate classes.

After identifying the optimal LCA solution, individuals were assigned to the latent class for which they had the highest posterior probability. Conditional item response probabilities were used to investigate within cluster prevalence of each individual LTC, then the clusters were subsequently labelled based on within and between cluster prevalence of these LTCs, as detailed below.

### Applied convention for labelling MLTC clusters

- Considered between cluster differences first, include LTCs that had substantially higher prevalence in one cluster compared to all others in name (rule of thumb approximately twice than next highest cluster)
- Labels include LTCs with 100% within-cluster prevalence and/or the highest between-cluster prevalence
- Labelled according to affected systems where LTCs with highest within and between prevalence affect similar physiology and body systems (e.g. Pulmonary, cardiovascular, etc.)
- Used ‘+’ when cluster named after single condition with highest within and/or between cluster prevalence
- Where a single LTC dominates the cluster, a + was added to the name to indicate that a collection of other LTCs are also in this cluster albeit without other obvious between-cluster differences that warrant naming
- Clusters with several LTCs with high within and between cluster prevalence which affect multiple systems and without one single dominant LTC in the cluster, were labelled Discordant

### Note

The clusters have been named for convenience in discussion and write-up. Clusters are complex and no naming convention will comprehensively describe a cluster. Readers are advised to investigate the prevalence of LTCs in detail when interpreting the data and results.

## Results

### Identified MLTC clusters

Below we present elbow plots containing model fit criteria and statistics and rationale for selection of the optimal LCA model in each age group and dataset combination.

#### SAIL LCA Models

**Figure 1.**
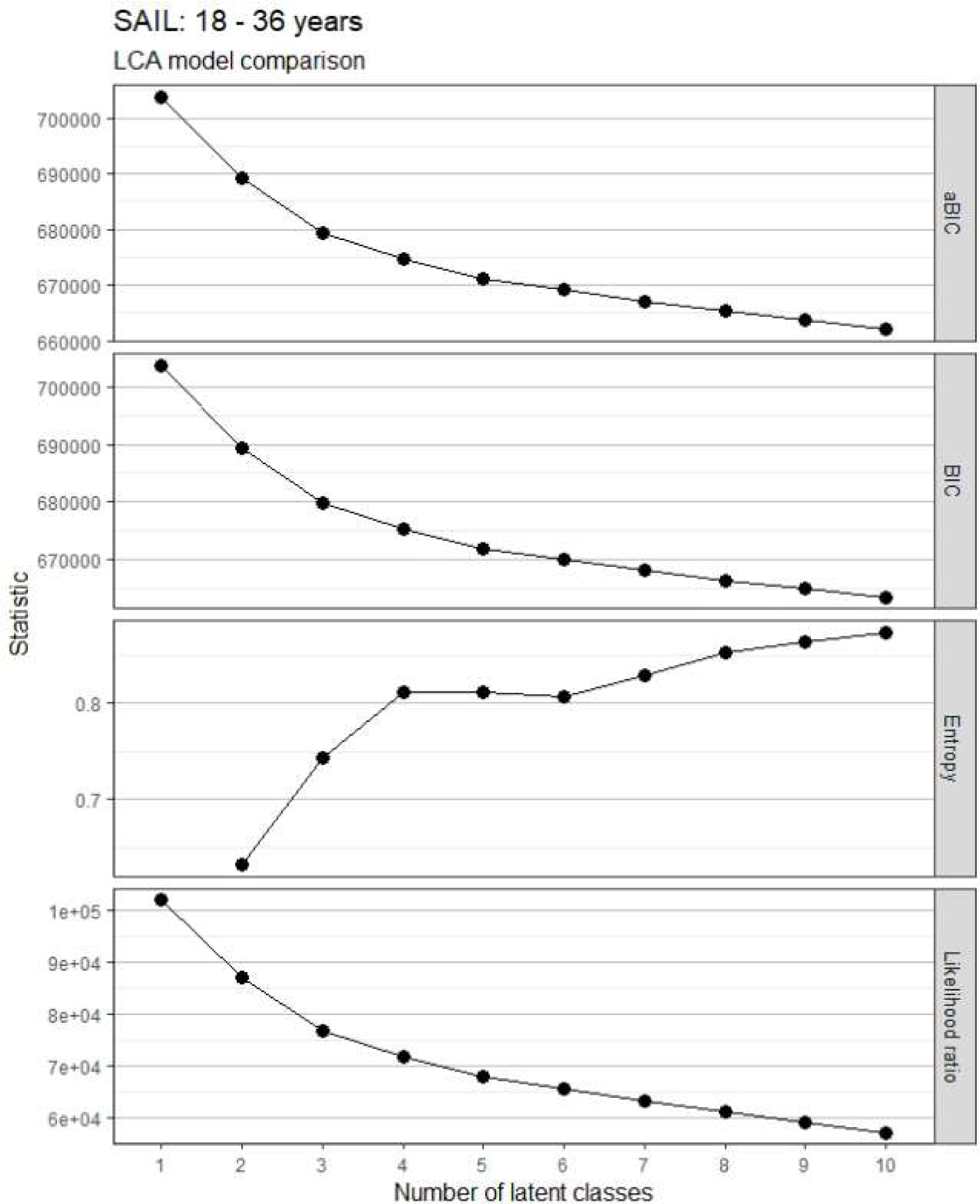
Model fit statistics for LCA models in adults 18-36 years in SAIL.

Model selected: 5-class model.

Reason: Stepwise improvement in aBIC and BIC was reduced beyond the 5-class solution. Only local maxima were identified in 6-class model and beyond, meaning poor data fit and models being rejected. 5-class model had good clinical interpretability and better fit statistics than 4-class model, with almost identical classification.

**Figure 2.**
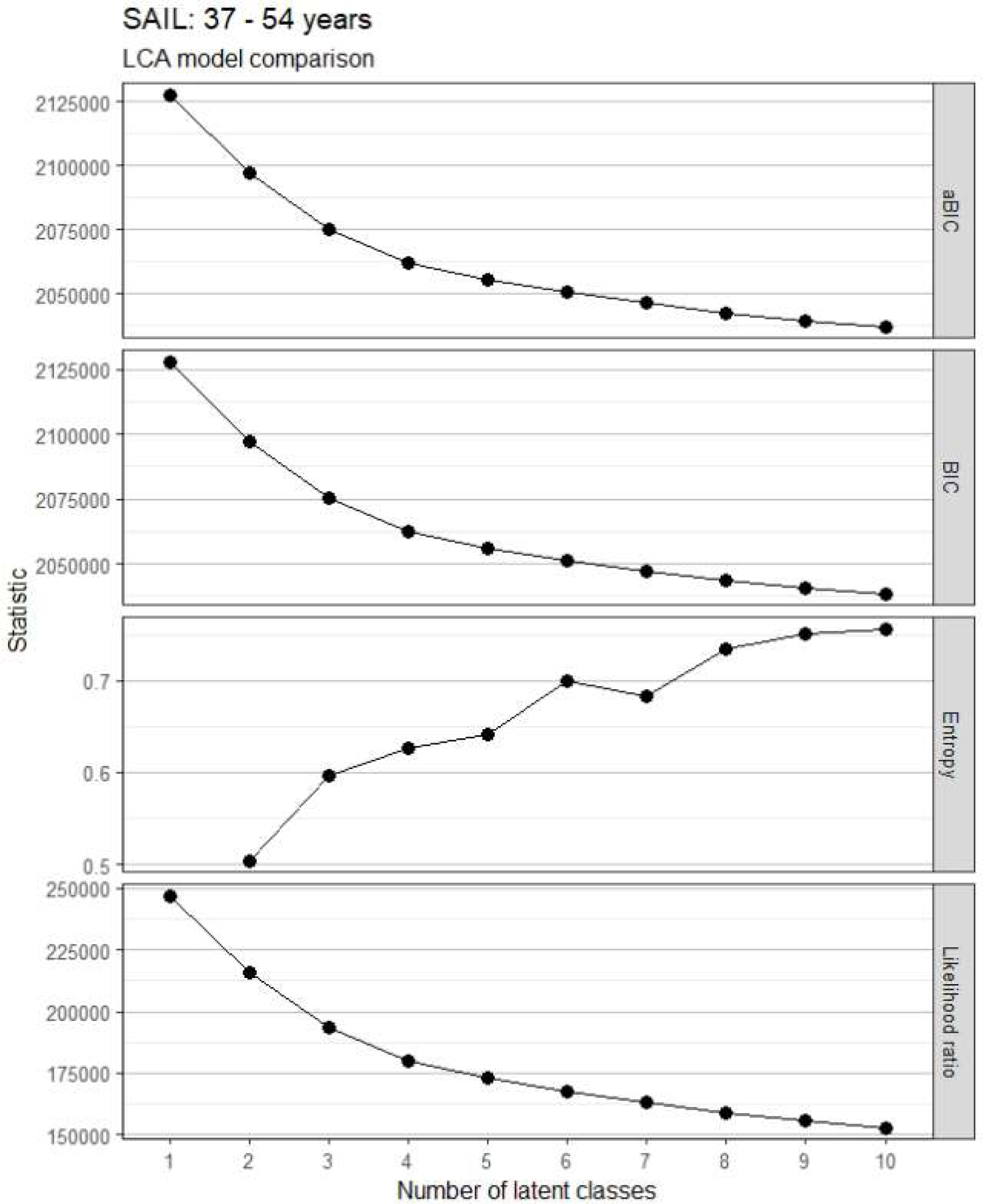
Model fit statistics for LCA models in adults 37-54 years in SAIL.

Model selected: 5-class model.

Rationale: Stepwise improvement in aBIC and BIC reduces beyond the 5-class model. Failure to replicate maximum likelihood function in 7-class model onwards. Marginal improvement in model classification between 5-class and 6-class model, but 6-class did not improve clinical interpretability.

**Figure 3.**
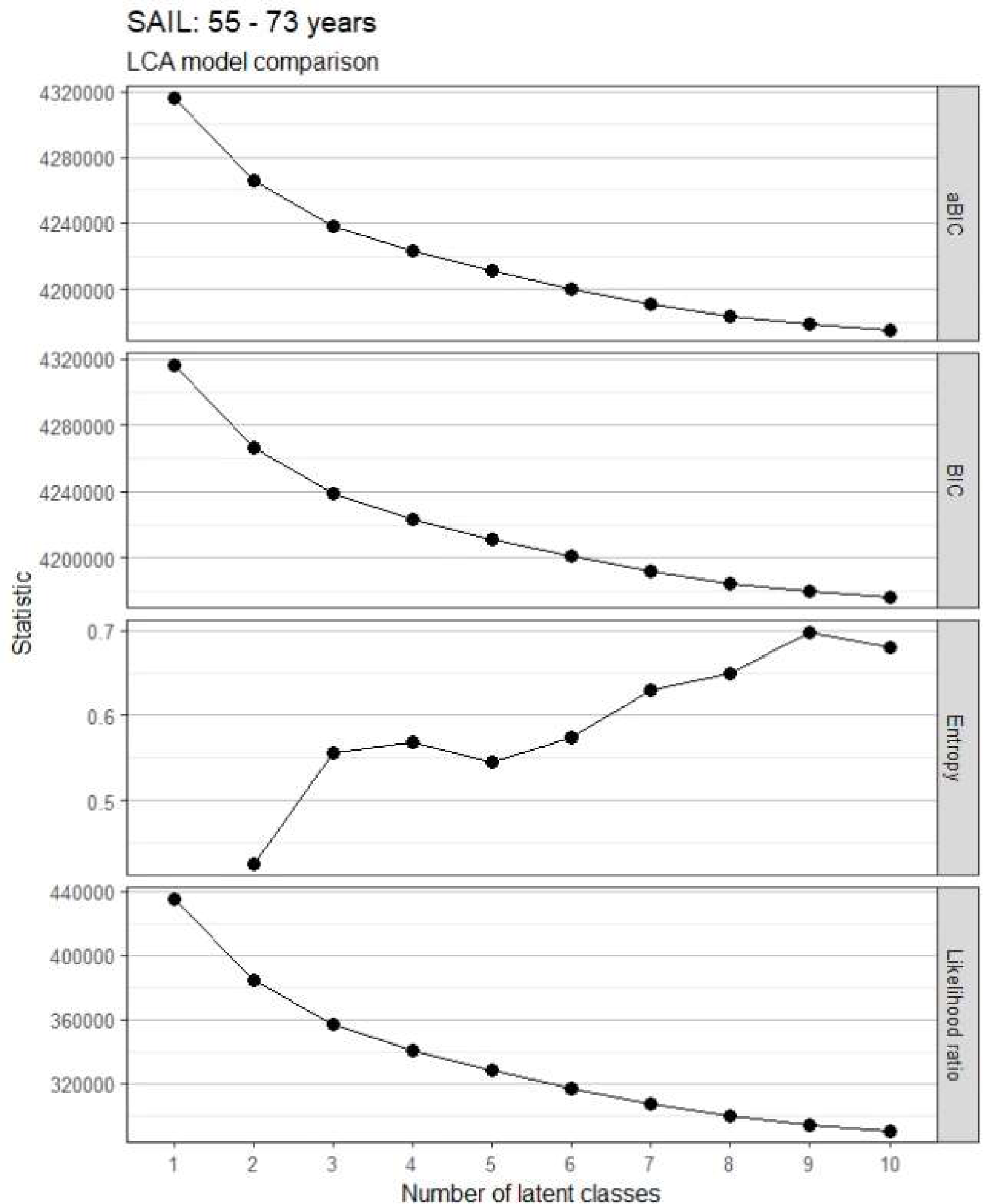
Model fit statistics for LCA models in adults 55 - 73 years in SAIL.

Model selected: 7-class model.

Rationale: Improvement in aBIC and BIC smaller after 6-class model. Classification improves in 7-class model. Clinical interpretation of 7-class model better than that of additional models with small differences in fit criteria and classification.

**Figure 4.**
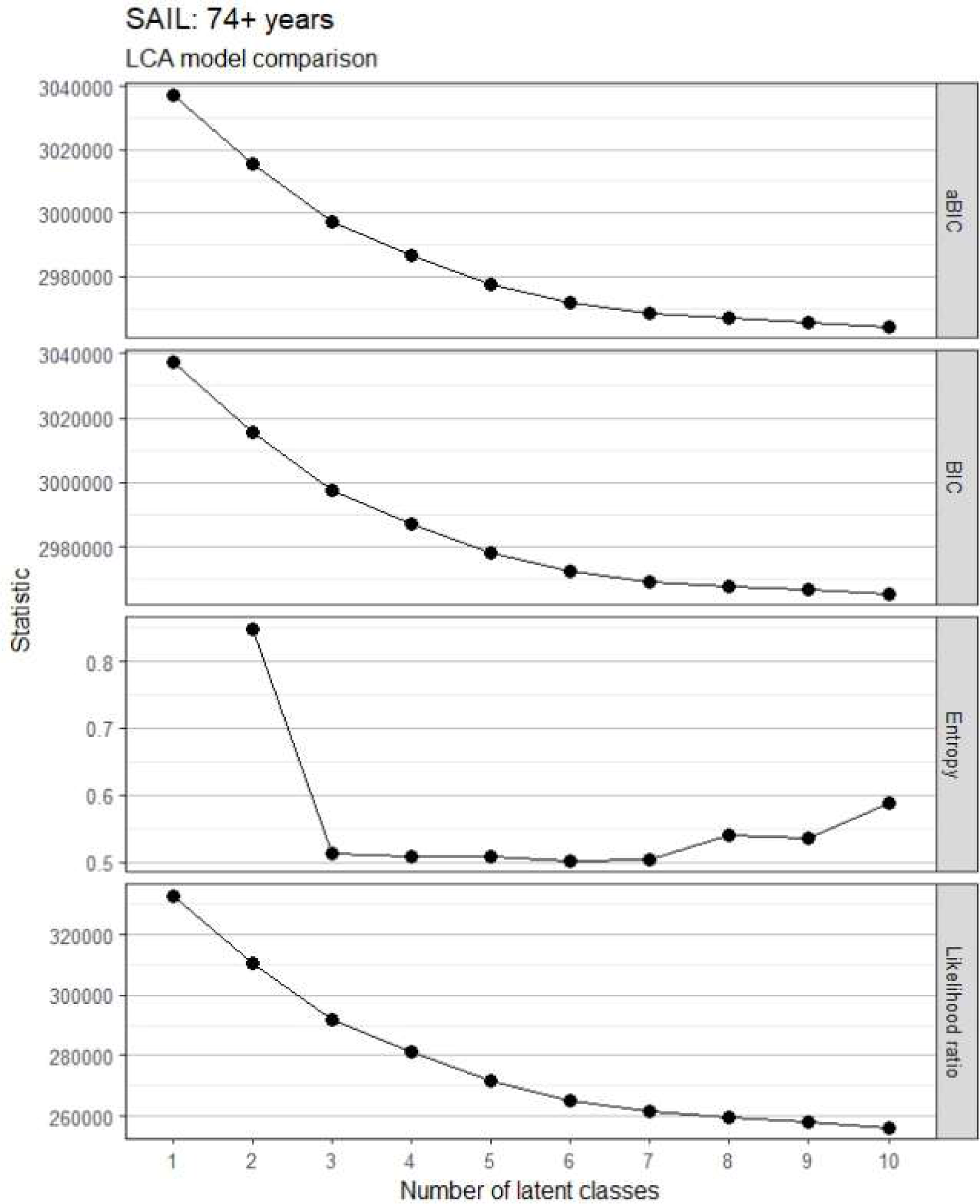
Model fit statistics for LCA models in adults 55 - 73 years in SAIL.

Model selected: 6-class model.

Rationale: Stepwise improvement in aBIC and BIC reduces around 5 to 6-class model. All models beyond 2-class model have similar classification. Two class model not appropriate as showed very similar clinical profiles split only by the presence/absence of COPD. Similar model fit between 6-class and 7-class, but clinical interpretation favourable in 6-class model.

#### UK Biobank LCA Models

**Figure 5.**
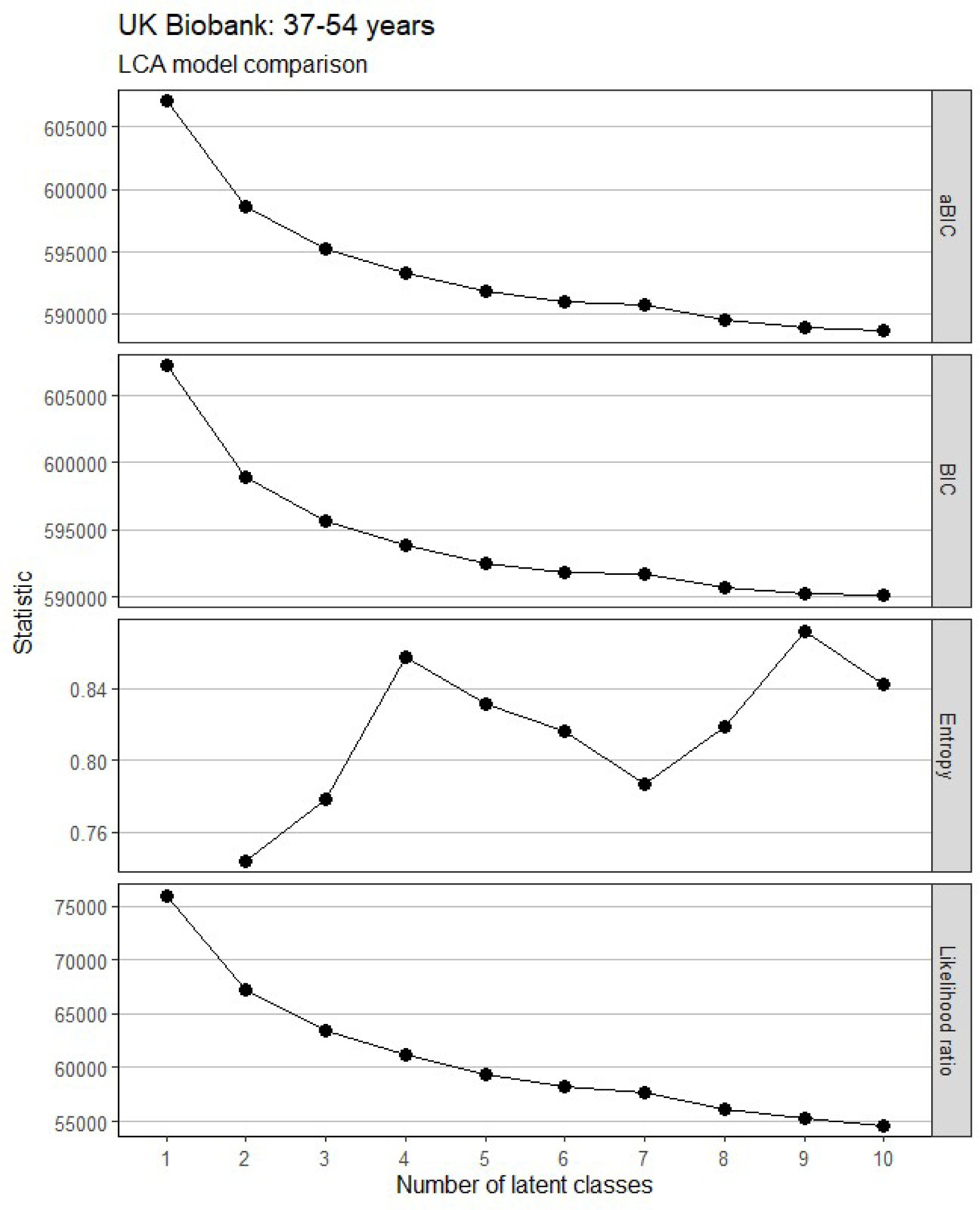
Model fit statistics for LCA models in adults 37 – 54 years in UK Biobank.

Model selected: 5-class model.

Rationale: Stepwise improvement in aBIC and BIC reduced beyond the 5-class model. Failed to replicate maximum log-likelihood function in 6-class model onwards, so models rejected due to instability and poor data fit.

**Figure 6.**
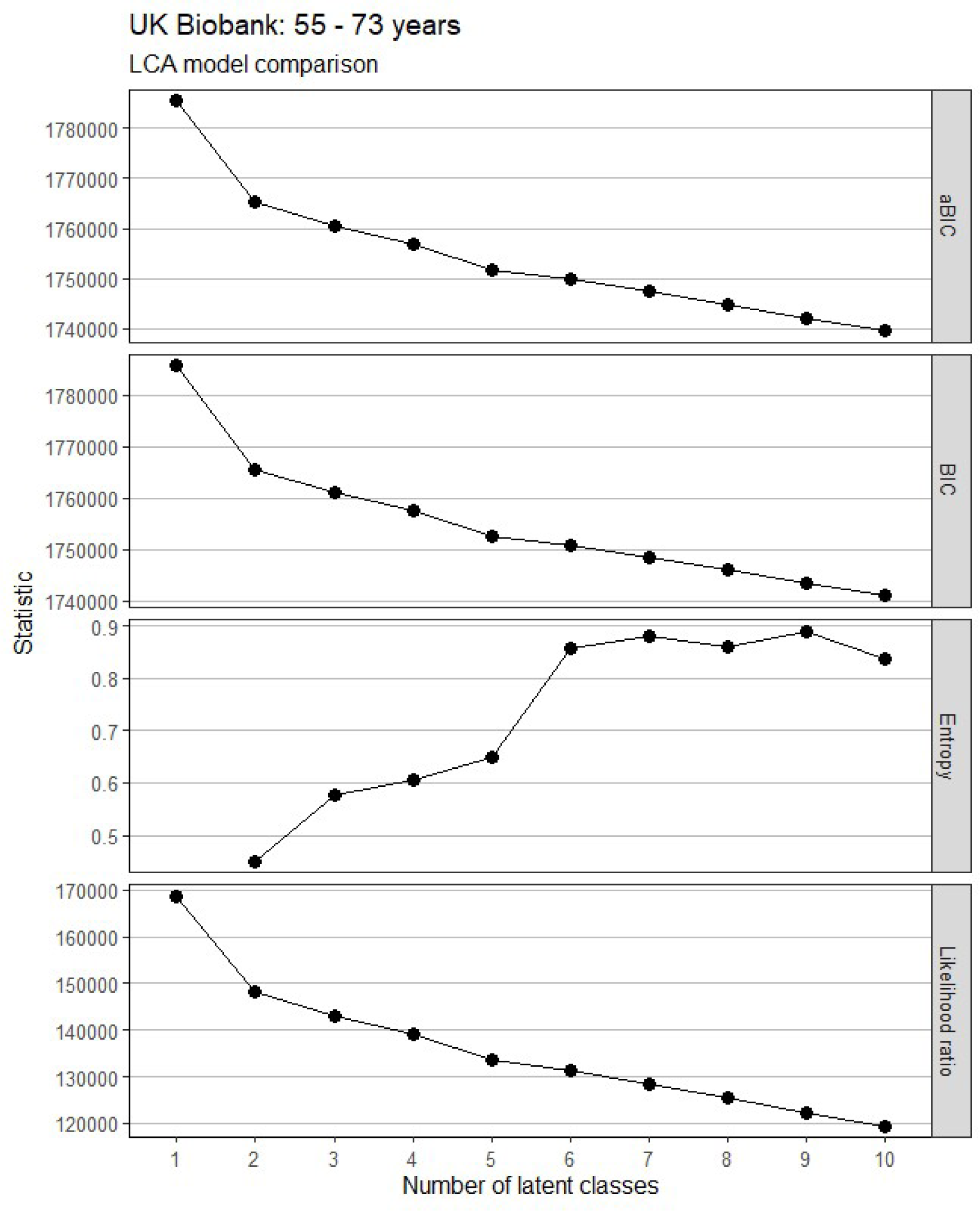
Model fit statistics for LCA models in adults 55 - 73 years in UK Biobank.

Model selected: 4-class model.

Rationale: Stepwise improvement in aBIC and BIC reduced beyond the 5-class model. However, failed to replicate maximum log-likelihood function in 5-class model onwards, so these models were rejected due to instability and poor data fit.

#### UKHLS LCA Models

**Figure 7.**
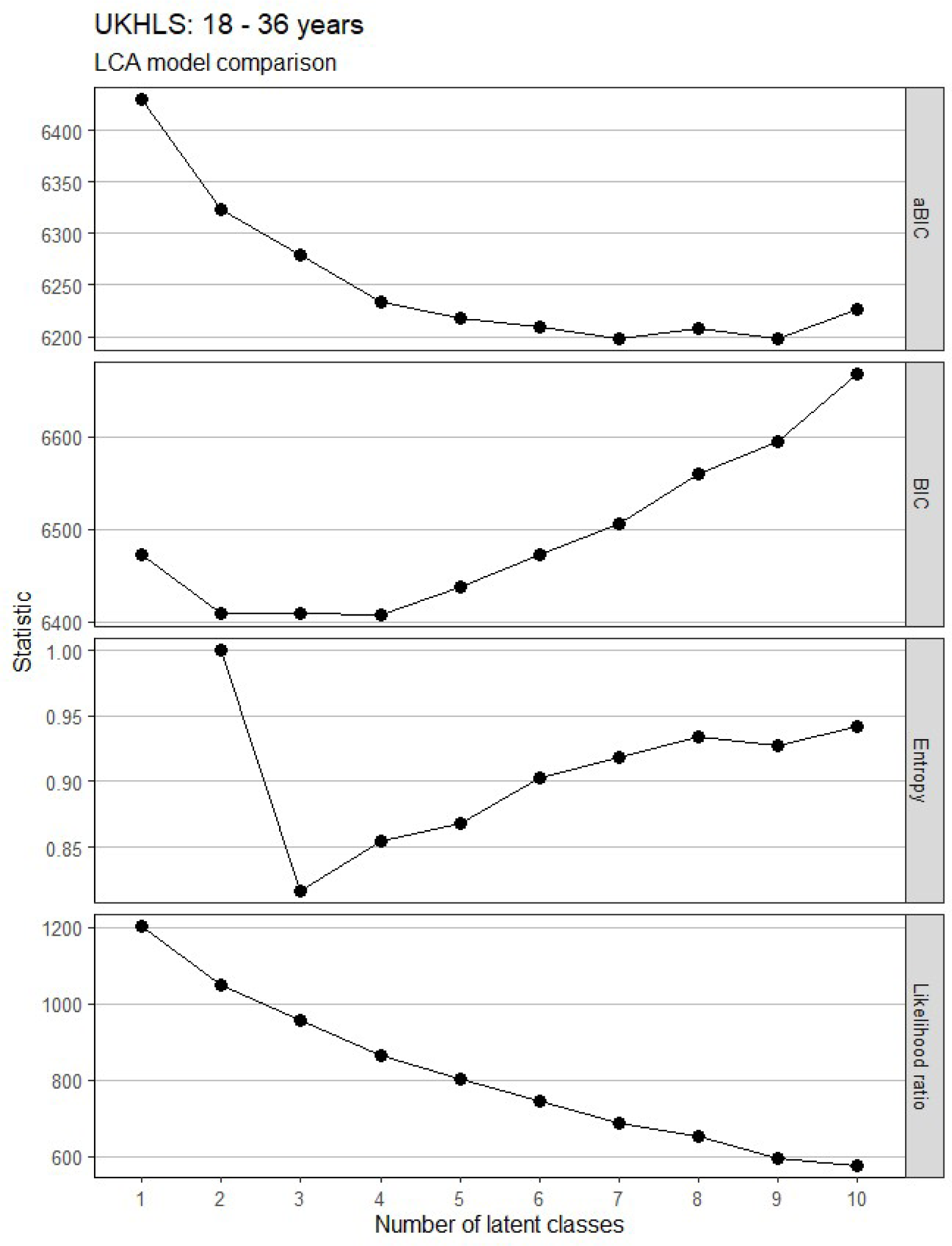
Model fit statistics for LCA models in adults 18 – 36 years in UKHLS.

Model selected: 3-class model.

Rationale: BIC almost identical in 2, 3 and 4-class models, reduction in aBIC improvement beyond 4-class model. Failed to replicate maximum log-likelihood in 4-class models and beyond so these were rejected. Clinical interpretation of 3-class model preferable vs 2-class model. Classification of this model was still good despite being the lowest among comparable models.

**Figure 8.**
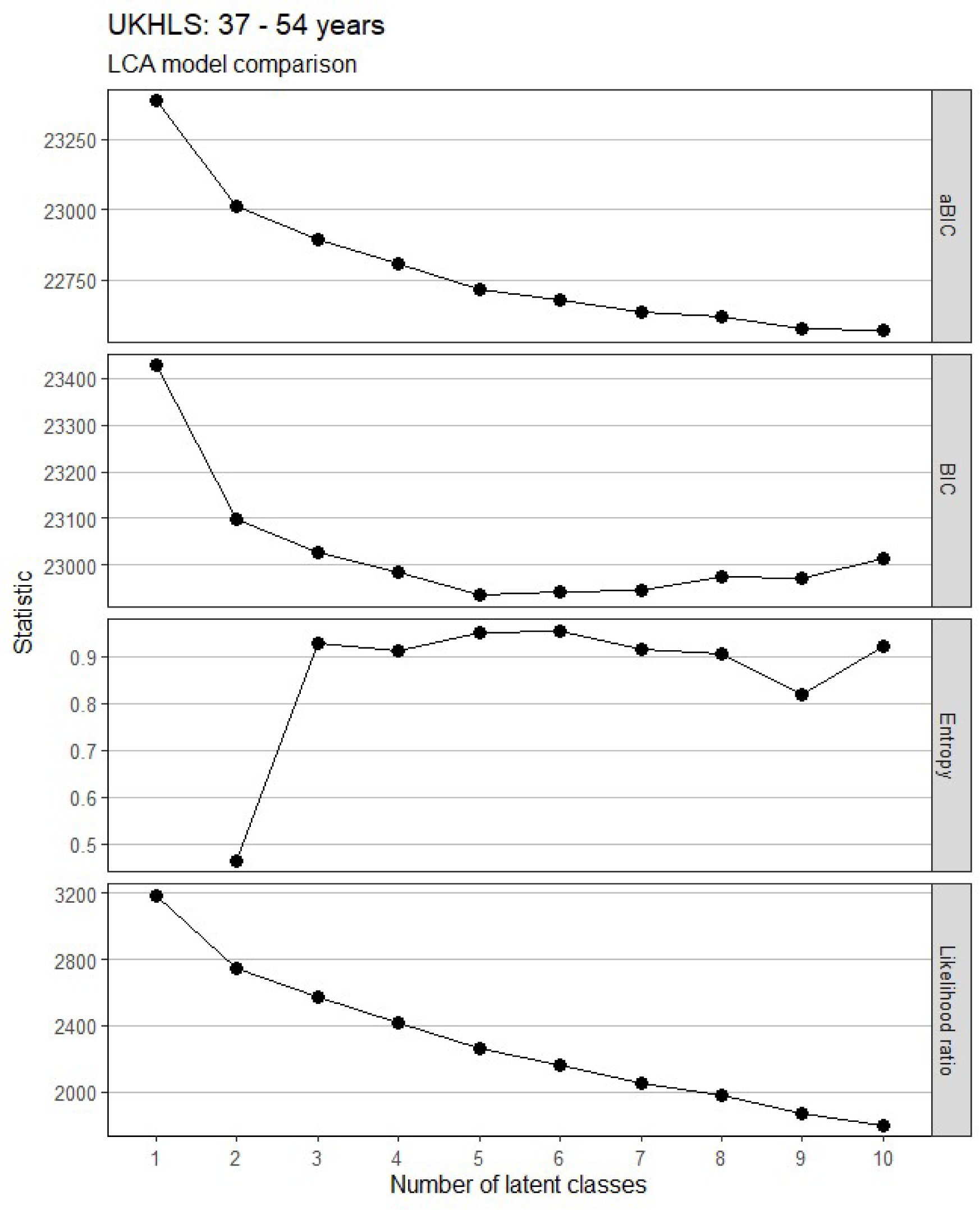
Model fit statistics for LCA models in adults 37 - 54 years in UKHLS.

Model selected: 5-class model.

Rationale: Lowest BIC and deflection of aBIC at 5-class model. Larger models were unstable and therefore rejected. Very clear clinical interpretation.

**Figure 9.**
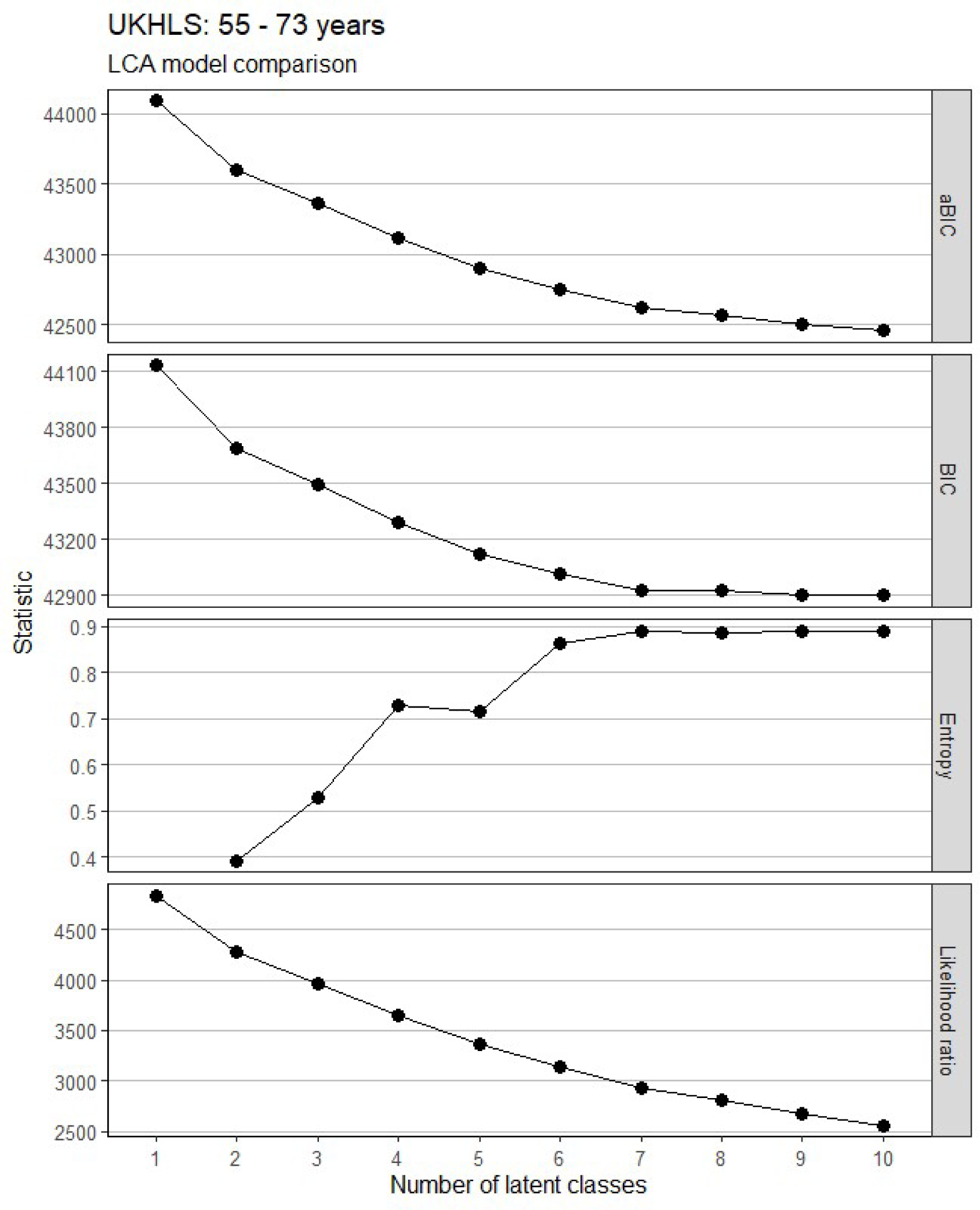
Model fit statistics for LCA models in adults 55 – 73 years in UKHLS.

Model selected: 4-class model.

Rationale: BIC and aBIC showed consistent improvement until 7-class model, but maximum log-likelihood function could not be replicated in any models beyond the 4-class model. Indeed, the EM algorithm failed to converge after 5000 iterations in the 7 to 10-class models, highlighting instability of these models. 4-class model had improved classification and clinical interpretation compared to smaller models.

**Figure 10.**
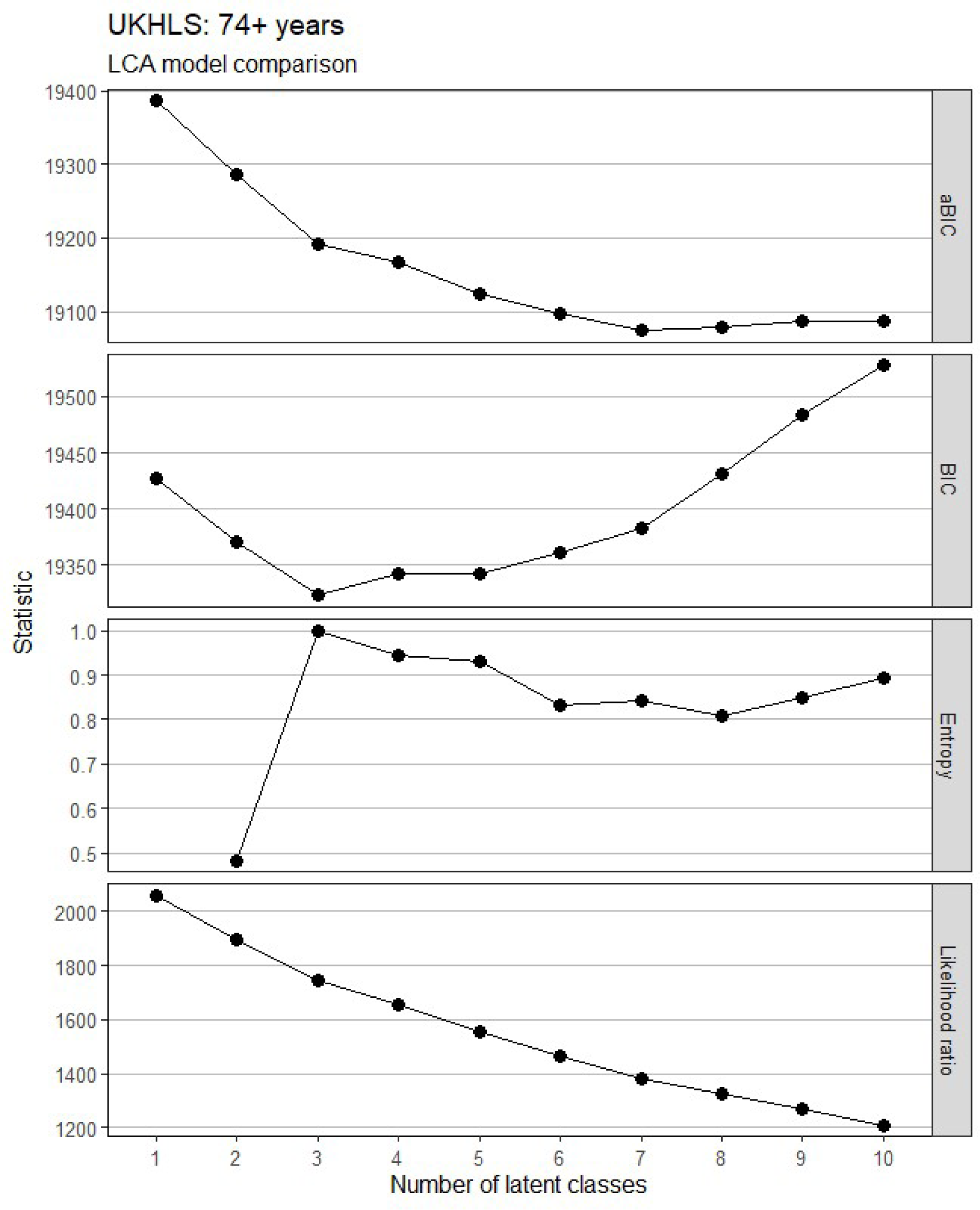
Model fit statistics for LCA models in adults 74+ years in UKHLS.

Model selected: 3-class model.

Rationale: Lowest BIC and reduced improvement of aBIC beyond the 3-class model. Model classification in 3-class model is ideal (entropy = 1). Model failed to converge in 4-class and 6-class model, and maximum likelihood function could not be replicated in 7-class model onwards.

### MLTC clusters

#### SAIL MLTC Clusters

**Figure 11.**
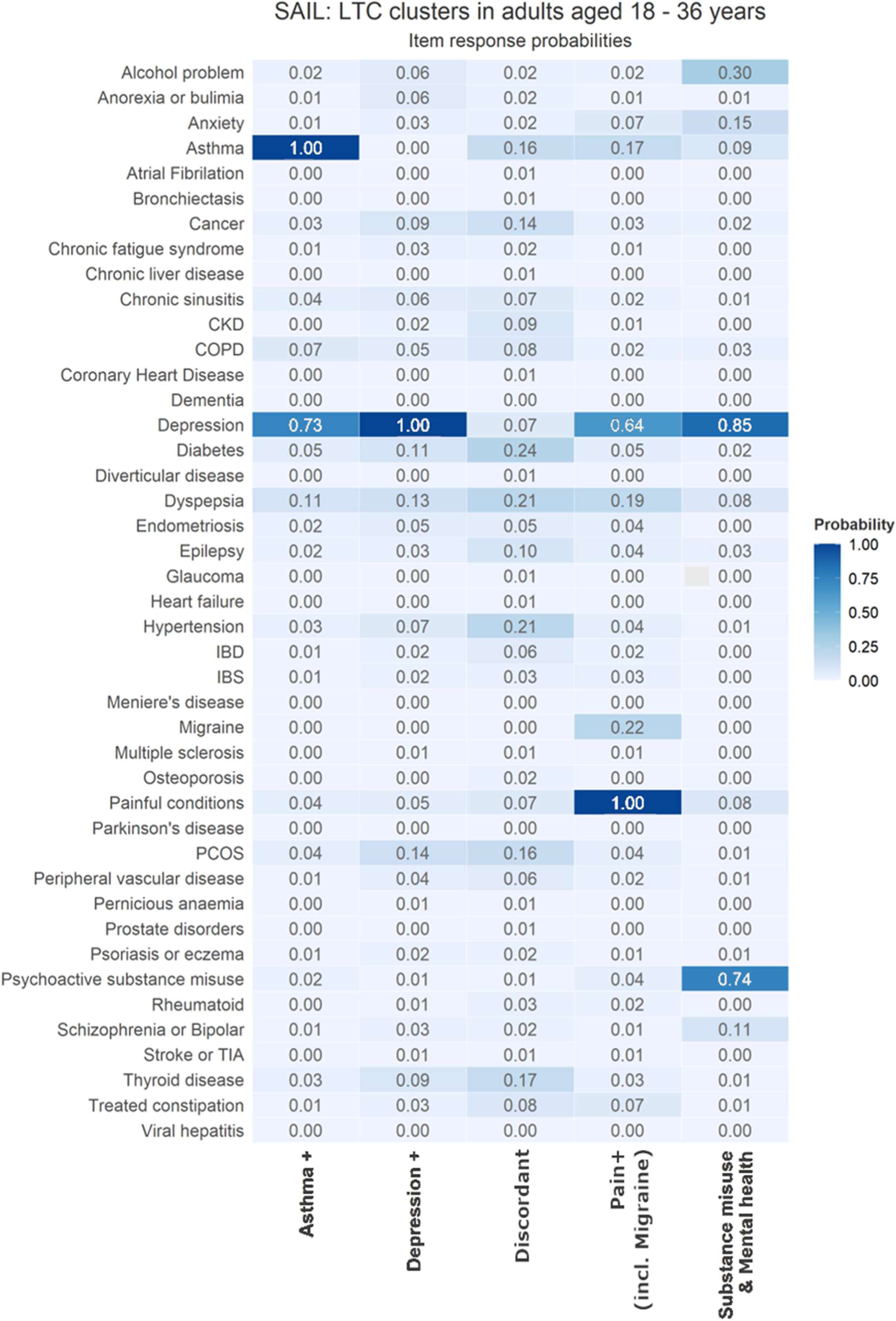
Within cluster conditional item response probabilities for adults 18 - 36 years in SAIL.

**Figure 12.**
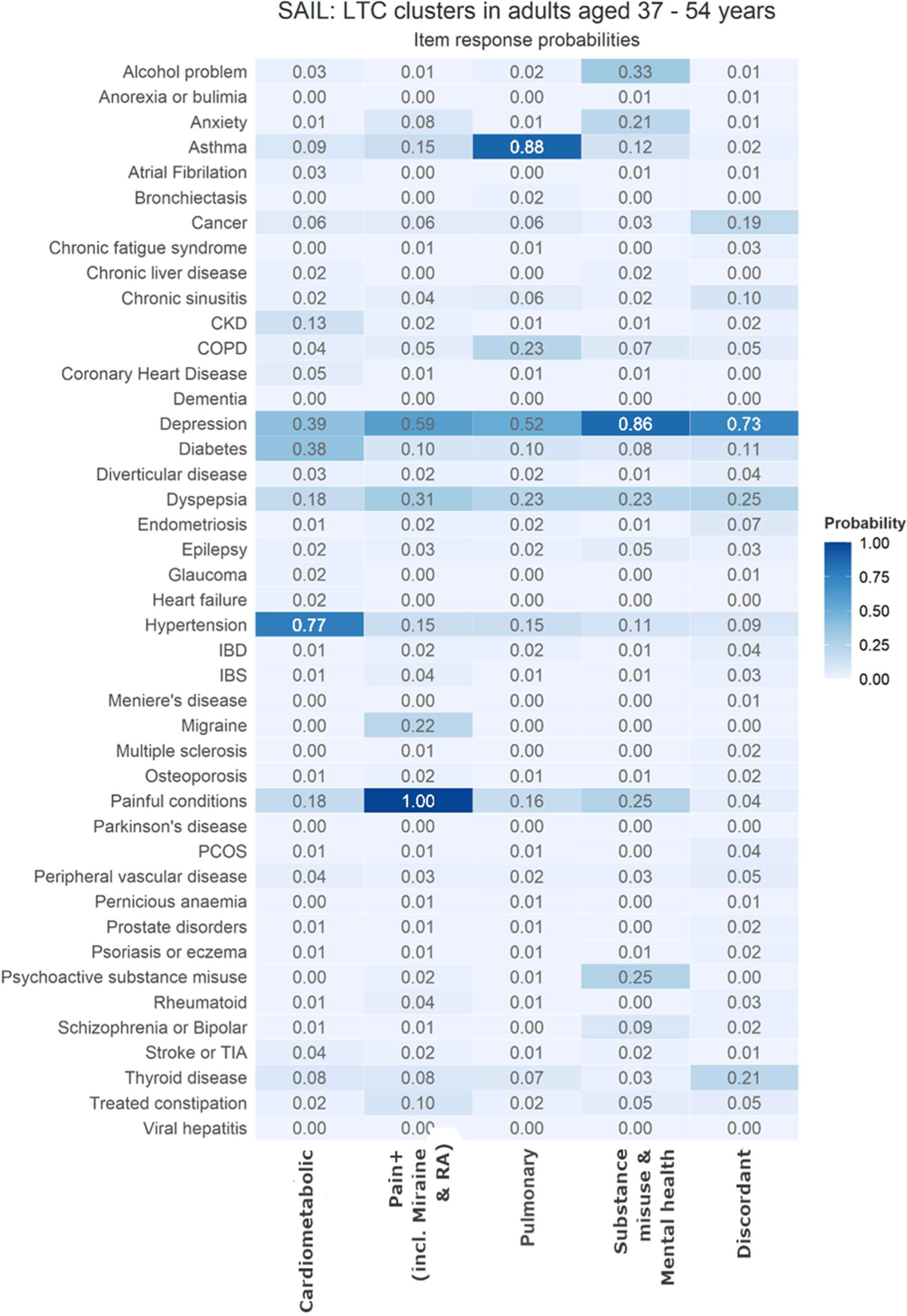
Within cluster conditional item response probabilities for adults 37 - 54 years in SAIL.

**Figure 13.**
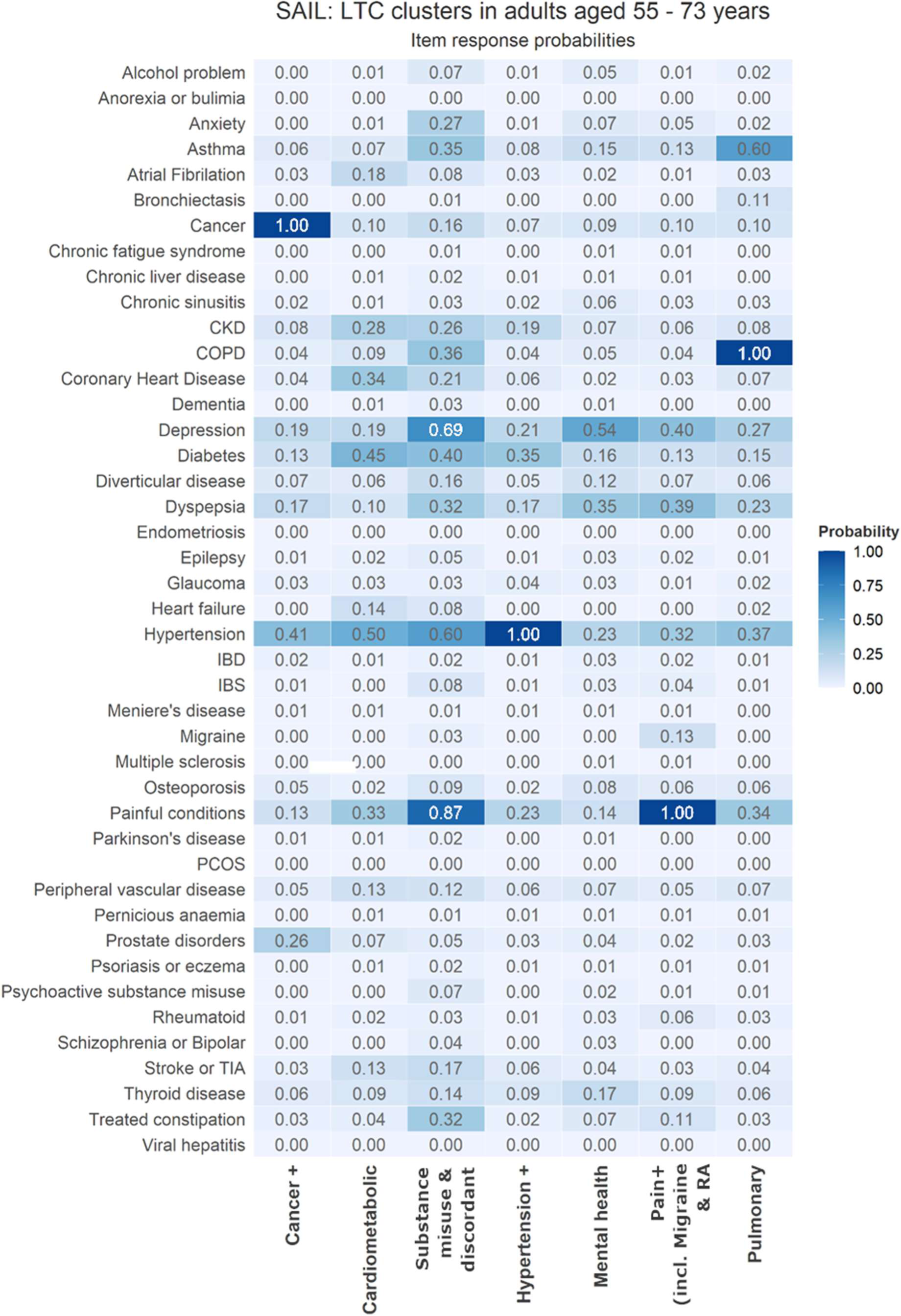
Within cluster conditional item response probabilities for adults 55 - 73 years in SAIL.

**Figure 14.**
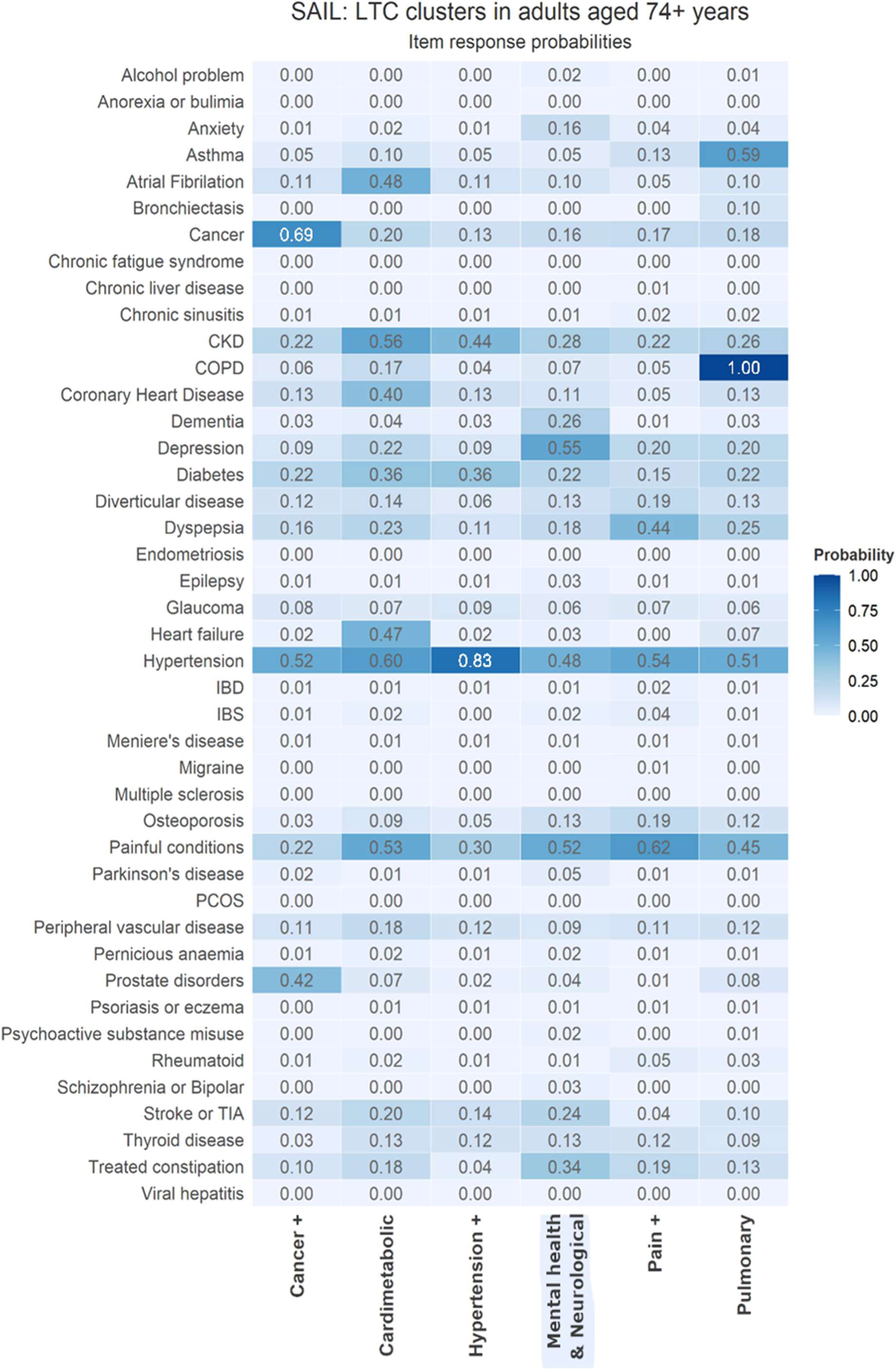
Within cluster conditional item response probabilities for adults 74+ years in SAIL.

### UK Biobank MLTC clusters

**Figure 15.**
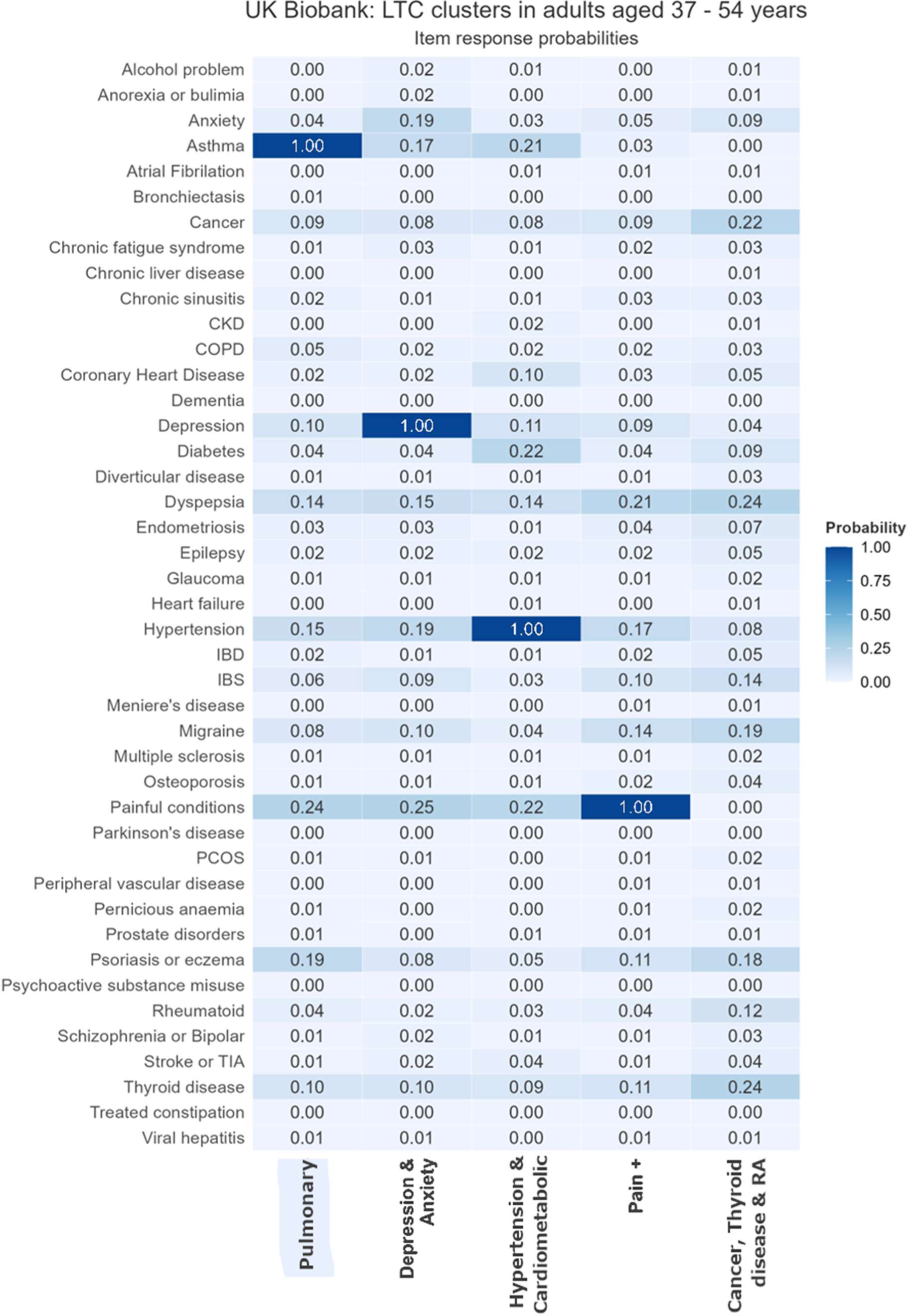
Within cluster conditional item response probabilities for adults 37 - 54 years in UK Biobank.

**Figure 16.**
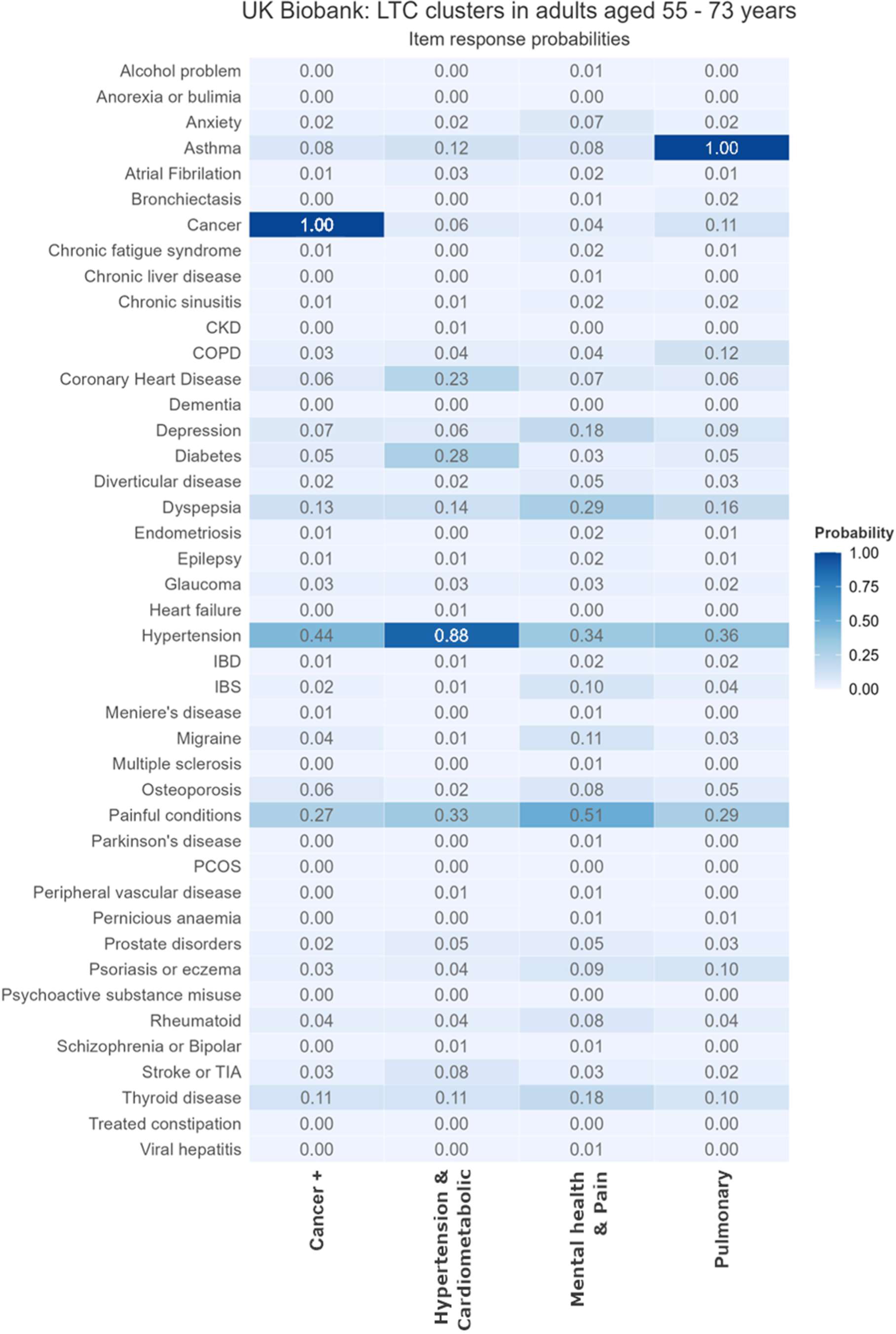
Within cluster conditional item response probabilities for adults 55-73 years in UK Biobank.

### UKHLS MLTC clusters

**Figure 17.**
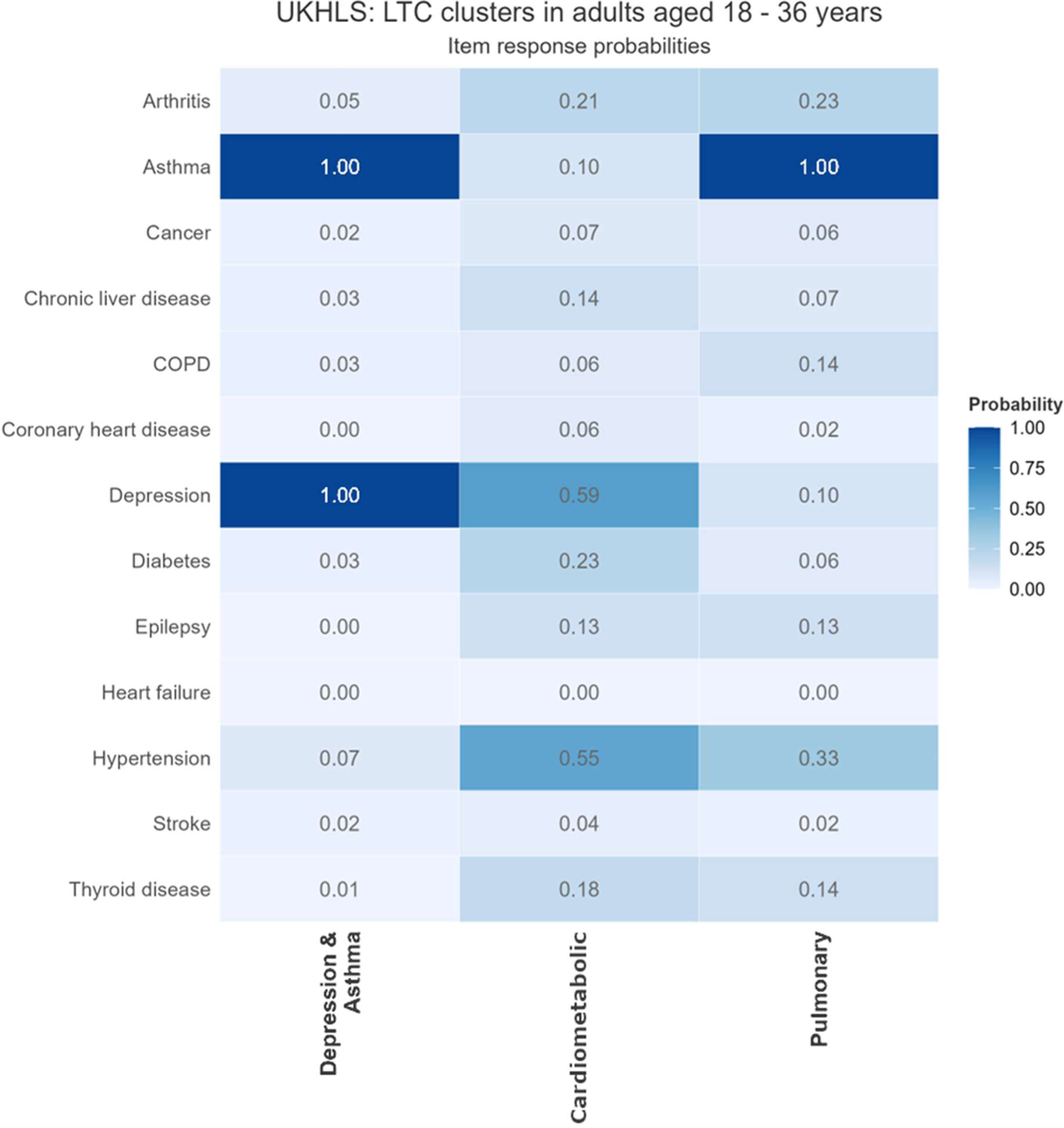
Within cluster conditional item response probabilities for adults 18 - 36 years in UKHLS.

**Figure 18.**
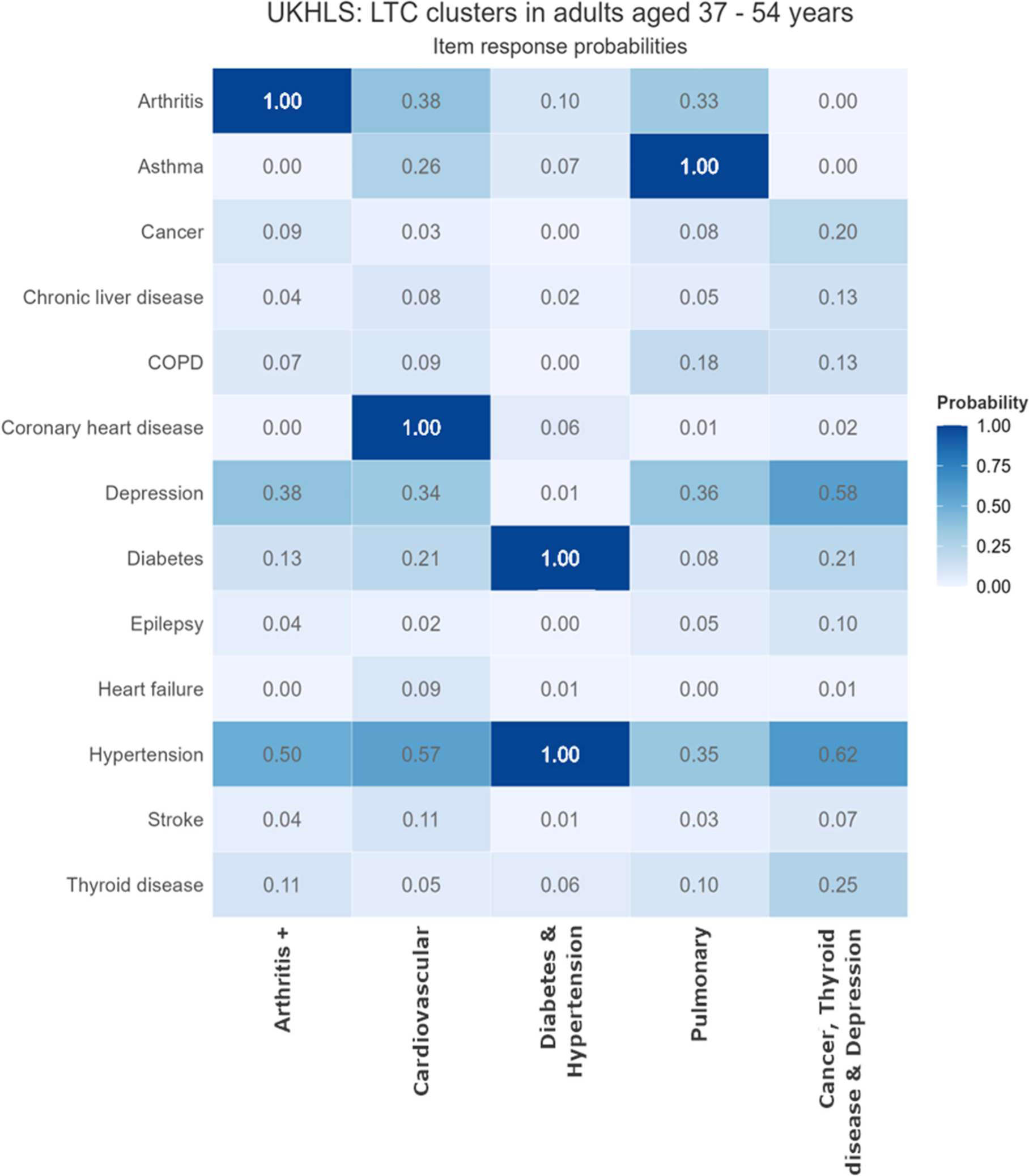
Within cluster conditional item response probabilities for adults 37 – 54 years in UKHLS.

**Figure 19.**
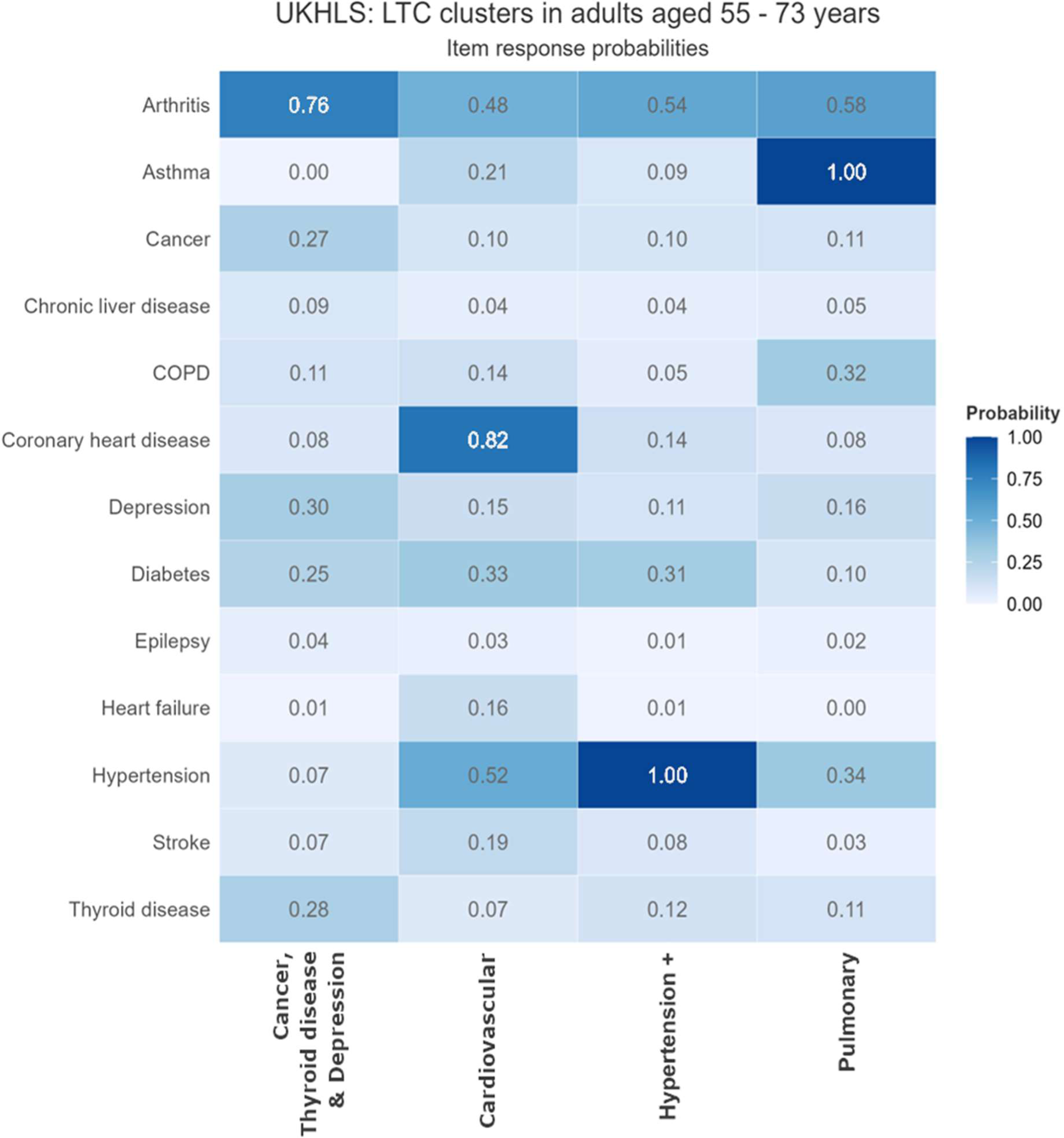
Within cluster conditional item response probabilities for adults 55 - 73 years in UKHLS.

**Figure 20.**
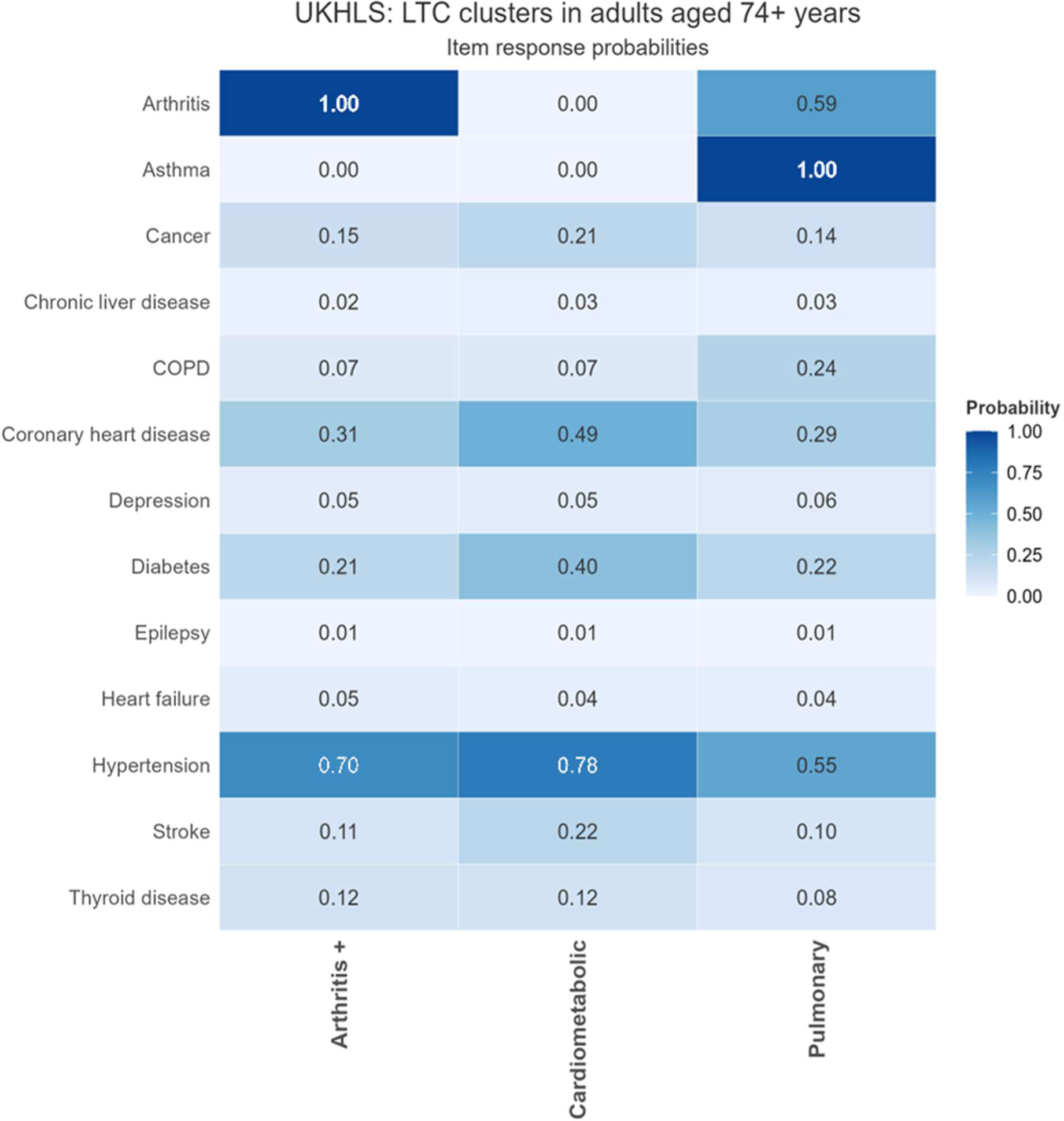
Within cluster conditional item response probabilities for adults 74+ years in UKHLS.

### MLTC Clusters – Sociodemographic characteristics

#### SAIL

##### SAIL: 18-36 years

**Table 2.** Sociodemographic characteristics of adults 18-36 years in SAIL stratified by multimorbidity clusters.

|  | No multimorbidity | Asthma+ | Pain+ (incl. Migraine) | Substance misuse & Mental health | Depression+ | Discordant |
| --- | --- | --- | --- | --- | --- | --- |
| N (%) | 503,188 (90.3) | 8,085 (1.5) | 10,312 (1.9) | 11,935 (2.1) | 17,698 (3.2) | 5,779 (1.0) |
| Female, n (%) | 235,864 (46.9) | 5,240 (64.8) | 7,280 (70.5) | 4,425 (37.1) | 13,445 (76.0) | 3,597 (62.2) |
| Age (SD) | 26.9 (5.5) | 28.9 (5.2) | 30.1 (4.9) | 29.5 (4.7) | 29.8 (4.8) | 29.9 (5.1) |
| Number of LTCs<br>[long list n=43] | 0.25 (0.43) | 2.33 (0.66) | 2.87 (1.10) | 2.61 (0.95) | 2.30 (0.63) | 2.35 (0.79) |
| Smoking, n (%) |  |  |  |  |  |  |
| Never | 299,015 (63.4) | 4,155 (51.4) | 4,746 (46.1) | 1,531 (13.0) | 8,394 (47.6) | 3,829 (66.6) |
| Previous | 134,116 (28.4) | 3,164 (39.1) | 4,377 (42.5) | 7,847 (66.6) | 7,464 (42.4) | 1,615 (28.1) |
| Current | 38,487 (8.2) | 764 (9.5) | 1,168 (11.4) | 2,402 (20.4) | 1,765 (10.0) | 306 (5.3) |
| Deprivation score<sup>1</sup> (SD) | 23.32 (15.79) | 25.41 (16.27) | 28.74 (17.30) | 32.10 (17.94) | 25.82 (16.42) | 23.86 (15.86) |
| Deprivation rank<sup>2</sup> (SD)<br>[median] | 904.1 (557.9)<br>[875] | 824.3 (550.4)<br>[752] | 715.5 (528.6)<br>[621] | 614.2 (511.0)<br>[478] | 810.8 (548.4)<br>[739] | 881.3 (553.0)<br>[851] |

<sup>1</sup> Welsh Government measure of relative multiple deprivation for small areas. Deprivation score (higher score=higher level of deprivation) rated across 8 types of deprivation employment, income, education, health, community safety, geographical access to services, housing, physical environment. <sup>2</sup> Welsh Government measure of relative multiple deprivation for small areas – relative ranking from 1 (most deprived area) to 1896 (least deprived area)

### SAIL: adults 37 – 54 years

**Table 3.** Sociodemographic characteristics of adults 37 – 54 years in SAIL stratified by multimorbidity cluster.

|  | No multimorbidity | Pulmonary | Pain+ (incl. Migraine & RA) | Substance misuse & Mental health | Cardiometabolic | Discordant |
| --- | --- | --- | --- | --- | --- | --- |
| N (%) | 439,997 (75.9) | 15,873 (2.7) | 36,929 (6.4) | 18,740 (3.2) | 34,804 (6.0) | 33,597 (5.8) |
| Female, n (%) | 203,289 (46.2) | 9,567 (60.3) | 25,275 (68.4) | 7,841 (41.8) | 15,846 (45.5) | 23,833 (70.9) |
| Age (SD) | 45.62 (5.04) | 46.52 (5.08) | 47.12 (4.96) | 45.39 (5.05) | 48.60 (4.67) | 46.20 (5.09) |
| Number of LTCs [long list n=43] | 0.36 (0.48) | 2.71 (1.02) | 3.25 (1.32) | 3.23 (1.40) | 2.77 (1.12) | 2.30 (0.64) |
| Smoking, n (%) |  |  |  |  |  |  |
| Never | 235,696 (58.1) | 7,561 (47.7) | 16,199 (44.3) | 3,879 (21.0) | 19,111 (55.10) | 17,372 (52.4) |
| Previous | 144,061 (35.5) | 6,998 (44.1) | 17,011 (46.5) | 11,283 (61.0) | 13,380 (38.6) | 13,443 (40.5) |
| Current | 25,750 (6.4) | 1,298 (8.2) | 3,398 (9.3) | 3,350 (18.1) | 2,194 (6.3) | 2,360 (7.1) |
| Deprivation score <sup>1</sup> (SD) | 20.85 (14.52) | 23.91 (15.96) | 27.06 (16.76) | 29.39 (17.53) | 24.26 (15.91) | 22.12 (14.97) |
| Deprivation rank <sup>2</sup> (SD) [median] | 992.8 (551.0) [991] | 881.6 (557.6) [844] | 767.2 (538.0) [683] | 697.0 (531.5) [586] | 864.4 (550.6) [818] | 941.6 (549.9) [924] |

<sup>1</sup> Welsh Government measure of relative multiple deprivation for small areas. Deprivation score (higher score=higher level of deprivation) rated across 8 types of deprivation employment, income, education, health, community safety, geographical access to services, housing, physical environment. <sup>2</sup> Welsh Government measure of relative multiple deprivation for small areas – relative ranking from 1 (most deprived area) to 1896 (least deprived area)

### SAIL: adults 55 – 73 years

**Table 4.**
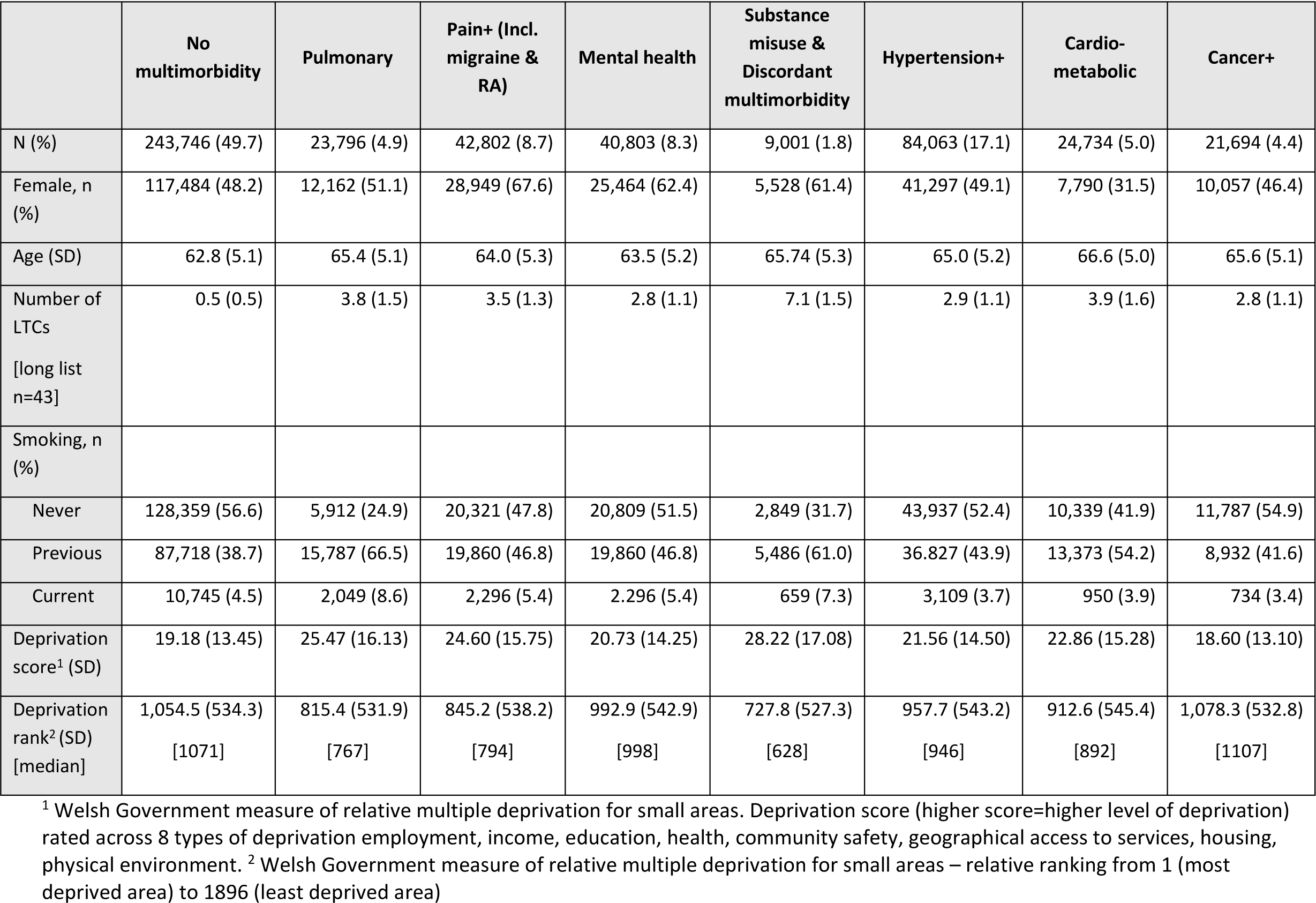
Sociodemographic characteristics of adults 55 - 73 years in SAIL stratified by multimorbidity cluster.

|  | No multimorbidity | Pulmonary | Pain+ (Incl. migraine & RA) | Mental health | Substance misuse & Discordant multimorbidity | Hypertension+ | Cardio-metabolic | Cancer+ |
| --- | --- | --- | --- | --- | --- | --- | --- | --- |
| N (%) | 243,746 (49.7) | 23,796 (4.9) | 42,802 (8.7) | 40,803 (8.3) | 9,001 (1.8) | 84,063 (17.1) | 24,734 (5.0) | 21,694 (4.4) |
| Female, n (%) | 117,484 (48.2) | 12,162 (51.1) | 28,949 (67.6) | 25,464 (62.4) | 5,528 (61.4) | 41,297 (49.1) | 7,790 (31.5) | 10,057 (46.4) |
| Age (SD) | 62.8 (5.1) | 65.4 (5.1) | 64.0 (5.3) | 63.5 (5.2) | 65.74 (5.3) | 65.0 (5.2) | 66.6 (5.0) | 65.6 (5.1) |
| Number of LTCs<br>[long list n=43] | 0.5 (0.5) | 3.8 (1.5) | 3.5 (1.3) | 2.8 (1.1) | 7.1 (1.5) | 2.9 (1.1) | 3.9 (1.6) | 2.8 (1.1) |
| Smoking, n (%) |  |  |  |  |  |  |  |  |
| Never | 128,359 (56.6) | 5,912 (24.9) | 20,321 (47.8) | 20,809 (51.5) | 2,849 (31.7) | 43,937 (52.4) | 10,339 (41.9) | 11,787 (54.9) |
| Previous | 87,718 (38.7) | 15,787 (66.5) | 19,860 (46.8) | 19,860 (46.8) | 5,486 (61.0) | 36,827 (43.9) | 13,373 (54.2) | 8,932 (41.6) |
| Current | 10,745 (4.5) | 2,049 (8.6) | 2,296 (5.4) | 2,296 (5.4) | 659 (7.3) | 3,109 (3.7) | 950 (3.9) | 734 (3.4) |
| Deprivation score<sup>1</sup> (SD) | 19.18 (13.45) | 25.47 (16.13) | 24.60 (15.75) | 20.73 (14.25) | 28.22 (17.08) | 21.56 (14.50) | 22.86 (15.28) | 18.60 (13.10) |
| Deprivation rank<sup>2</sup> (SD)<br>[median] | 1,054.5 (534.3)<br>[1071] | 815.4 (531.9)<br>[767] | 845.2 (538.2)<br>[794] | 992.9 (542.9)<br>[998] | 727.8 (527.3)<br>[628] | 957.7 (543.2)<br>[946] | 912.6 (545.4)<br>[892] | 1,078.3 (532.8)<br>[1107] |

<sup>1</sup> Welsh Government measure of relative multiple deprivation for small areas. Deprivation score (higher score=higher level of deprivation) rated across 8 types of deprivation employment, income, education, health, community safety, geographical access to services, housing, physical environment. <sup>2</sup> Welsh Government measure of relative multiple deprivation for small areas – relative ranking from 1 (most deprived area) to 1896 (least deprived area)

### SAIL: adults (74+ years)

**Table 5.** Sociodemographic characteristics of adults 74+ years in SAIL stratified by multimorbidity cluster.

|  | No multimorbidity | Pulmonary | Pain+ | Mental health & Neurological | Hypertension+ | Cardiometabolic | Cancer+ |
| --- | --- | --- | --- | --- | --- | --- | --- |
| N (%) | 44,799 (22.7) | 15,432 (7.8) | 28,832 (14.6) | 16,136 (8.2) | 61,886 (31.3) | 13,220 (6.7) | 17,399 (8.8) |
| Female, n (%) | 24,809 (55.4) | 8,216 (53.2) | 22,393 (77.7) | 11,837 (73.4) | 37,859 (61.2) | 7,139 (54.0) | 4,377 (25.2) |
| Age (SD) | 80.4 (5.3) | 80.7 (4.9) | 81.3 (5.4) | 83.3 (5.9) | 81.4 (5.3) | 82.8 (5.6) | 81.1 (5.2) |
| Number of LTCs [long list n=43] | 0.58 (0.49) | 5.01 (2.00) | 3.95 (1.58) | 5.02 (1.91) | 3.30 (1.27) | 6.15 (2.03) | 3.52 (1.52) |
| Smoking, n (%) |  |  |  |  |  |  |  |
| Never | 25,081 (63.9) | 5,449 (35.4) | 18,408 (64.4) | 10,068 (62.8) | 37,994 (61.6) | 7,350 (55.8) | 9,823 (56.9) |
| Previous | 13,265 (33.8) | 9,400 (61) | 9,744 (34.1) | 5,651 (35.3) | 22,762 (36.9) | 5,680 (43.1) | 7,198 (41.7) |
| Current | 897 (2.3) | 552 (3.6) | 450 (1.6) | 304 (1.9) | 927 (1.5) | 148 (1.1) | 249 (1.4) |
| Deprivation score<sup>1</sup> (SD) | 19.84 (13.90) | 23.43 (15.36) | 20.81 (14.05) | 21.58 (14.43) | 20.34 (13.78) | 21.44 (14.37) | 18.19 (12.91) |
| Deprivation rank<sup>2</sup> (SD) [median] | 1,030.5 (546.9) [1032] | 888.5 (543.1) [858] | 985.3 (544.1) [977] | 955.7 (546.6) [937] | 1,002.1 (539.3) [997] | 960.8 (542.7) [954] | 1,095.2 (533.0) [1120] |

<sup>1</sup> Welsh Government measure of relative multiple deprivation for small areas. Deprivation score (higher score=higher level of deprivation) rated across 8 types of deprivation employment, income, education, health, community safety, geographical access to services, housing, physical environment. <sup>2</sup> Welsh Government measure of relative multiple deprivation for small areas – relative ranking from 1 (most deprived area) to 1896 (least deprived area)

#### UK BIOBANK

### UK BIOBANK: adults 37 – 54 years

**Table 6.** Sociodemographic characteristics of adults 37 – 54 years in UK Biobank stratified by multimorbidity cluster.

|  | No multimorbidity | Pulmonary | Pain+ | Depression & Anxiety | Hypertension & Cardiometabolic | Cancer, Thyroid disease & Rheumatoid Arthritis |
| --- | --- | --- | --- | --- | --- | --- |
| N (%) | 151,975 (78.3) | 9,680 (5.0) | 5,072 (2.6) | 5,250 (2.7) | 16,126 (8.3) | 6,006 (3.1) |
| Female, n (%) | 82,942 (54.6) | 6,174 (63.8) | 3,255 (64.2) | 3,702 (70.5) | 7,592 (47.1) | 4,204 (70.0) |
| Age (SD) | 47.5 (4.2) | 47.7 (4.2) | 48.6 (4.1) | 47.9 (4.1) | 49.2 (3.9) | 48.4 (4.1) |
| Number of LTCs (SD)<br>[long list n=43] | 0.4 (0.5) | 2.6 (0.9) | 2.5 (0.8) | 2.8 (1.1) | 2.6 (0.9) | 2.2 (0.6) |
| Townsend score<br>[missing data for n=330] | -1.11 (3.16) | -0.67 (3.36) | -0.59 (3.33) | -0.15 (3.48) | -0.24 (3.47) | -0.71 (3.34) |
| Deprivation quintiles (Townsend), n (%)<br>[missing data for n=330] |  |  |  |  |  |  |
| 1 (Least deprived) | 28,754 (19.0) | 1,614 (16.7) | 788 (15.6) | 703 (13.4) | 2,276 (14.1) | 1,023 (17.1) |
| 2 | 28,303 (18.7) | 1,596 (16.5) | 833 (16.5) | 763 (14.6) | 2,429 (15.1) | 1,007 (16.8) |
| 3 | 29,515 (19.5) | 1,772 (18.4) | 939 (18.5) | 917 (17.5) | 2,787 (17.3) | 1,102 (18.4) |
| 4 | 32,141 (21.2) | 2,079 (21.5) | 1,095 (21.6) | 1,130 (21.6) | 3,534 (22.0) | 1,258 (21.0) |
| 5 (Most deprived) | 33,025 (21.8) | 2,598 (26.9) | 1,410 (27.8) | 1,722 (32.9) | 5,067 (31.5) | 1,608 (26.8) |
| White ethnicity, n (%)<br>[missing data for n=1,149] | 138,121 (91.4) | 8,958 (93.0) | 4,685 (92.8) | 4,945 (94.7) | 14,279 (89.1) | 5,580 (93.4) |
| BMI (SD) [missing data for n=1,225] | 26.7 (4.6) | 28.0 (5.6) | 28.1 (5.4) | 28.2 3 (6.0) | 30.9 (6.2) | 27.0 (5.2) |
| Smoking, n (%)<br>[missing for n=983] |  |  |  |  |  |  |
| Never | 93,293 (61.7) | 5,481 (56.9) | 2,753 (54.6) | 2,675 (51.2) | 8,783 (54.7) | 3,367 (56.3) |
| Previous | 39,106 (25.9) | 2,684 (27.9) | 1,432 (28.4) | 1,392 (26.6) | 4,792 (29.9) | 1,717 (28.7) |
| Current | 18,802 (12.4) | 1,465 (15.2) | 854 (17.0) | 1,161 (22.2) | 2,475 (15.4) | 894 (15.0) |
| Alcohol (weekly units)<br>[missing data for n=1,377] | 15.3 (19.0) | 13.0 (18.6) | 12.4 (20.4) | 13.0 (25.8) | 17.3 (25.4) | 10.8 (18.0) |
| Alcohol group, n (%)<br>[missing data for n=1,377] |  |  |  |  |  |  |
| Low risk (<15 units/week) | 92,848 (61.5) | 6,517 (67.8) | 3,532 (70.1) | 3,693 (71.1) | 9,828 (61.4) | 4,422 (74.2) |
| Increasing risk (14 – 35 units/week) | 48,130 (31.9) | 2,551 (26.5) | 1,225 (24.3) | 1,126 (21.7) | 4,516 (28.2) | 1,267 (21.3) |
| High risk (>35 units/week) | 9,953 (6.6) | 549 (5.7) | 281 (5.6) | 376 (7.2) | 1,653 (10.3) | 267 (4.5) |
| Physical Activity, n (%)<br>[missing data for n= 2,867] |  |  |  |  |  |  |

|  | No<br>multimorbidity | Pulmonary | Pain+ | Depression &<br>Anxiety | Hypertension &<br>Cardiometabolic | Cancer,<br>Thyroid<br>disease &<br>Rheumatoid<br>Arthritis |
| --- | --- | --- | --- | --- | --- | --- |
| High | 25,984 (17.4) | 1,175 (12.3) | 446 (8.9) | 355 (6.9) | 1,256 (7.9) | 544 (9.2) |
| Medium | 110,615 (73.9) | 7,113 (74.4) | 3,726 (74.4) | 3,799 (73.4) | 11,697 (73.8) | 4,570 (77.1) |
| Low | 4,390 (2.9) | 385 (4.0) | 237 (4.7) | 281 (5.4) | 793 (5.0) | 237 (4.0) |
| None | 8,725 (5.8) | 894 (9.3) | 601 (12.0) | 743 (14.4) | 2,101 (13.3) | 575 (9.7) |
| Frailty<sup>1</sup>, n (%)<br>[data missing for n=4,405] |  |  |  |  |  |  |
| Robust | 96,507 (64.9) | 4,713 (49.7) | 2,310 (46.6) | 1,812 (35.4) | 6,511 (41.6) | 2,904 (49.5) |
| Pre-frail | 50,005 (33.6) | 4,183 (44.1) | 2,195 (44.3) | 2,720 (53.2) | 7,757 (49.6) | 2,652 (45.2) |
| Frail | 2,128 (1.4) | 589 (6.2) | 448 (9.1) | 584 (11.4) | 1,372 (8.8) | 314 (5.4) |
| Self-rated health, n (%)<br>[data missing for n=1,575] |  |  |  |  |  |  |
| Excellent | 32,014 (21.2) | 748 (7.8) | 295 (5.9) | 239 (4.6) | 608 (3.8) | 449 (7.6) |
| Fair | 24,641(16.3) | 3,083 (32.1) | 1,710 (34.1) | 1,929 (37.3) | 6,269 (39.3) | 1,934 (32.6) |
| Good | 91,191 (60.5) | 4,720 (49.2) | 2,339 (46.6) | 1,984 (38.3) | 6,508 (40.8) | 2,903 (48.9) |
| Poor | 3,008 (2.0) | 1,052 (11.0) | 678 (13.5) | 1,023 (19.8) | 2,555 (16.0) | 654 (11.0) |

All data presented as mean (SD) unless otherwise stated. <sup>1</sup> Frailty according to the frailty phenotype.

### UK BIOBANK: adults 55 – 73 years

**Table 7.** Sociodemographic characteristics of adults 55 - 73 years in UK Biobank stratified by multimorbidity cluster.

|  | No multimorbidity | Pulmonary | Mental Health & Pain | Hypertension & Cardiometabolic | Cancer+ |
| --- | --- | --- | --- | --- | --- |
| N (%) | 185,265 (60.1) | 20,624 (6.7) | 33,165 (10.8) | 52,740 (17.1) | 16,460 (5.3) |
| Female, n (%) | 98,778 (53.3) | 12,520 (60.7) | 21,891 (66.0) | 21,982 (41.7) | 10,259 (62.3) |
| Age | 61.6 (4.0) | 62.1 (4.1) | 62.3 (4.1) | 63.1 (4.0) | 63.2 (4.0) |
| Number of LTCs (SD) [long list n=43] | 0.5 (0.5) | 2.8 (1.0) | 2.7 (1.1) | 2.7 (1.0) | 2.6 (0.8) |
| Townsend score [missing data for n=293] | -1.71 (2.86) | -1.29 (3.11) | -1.31 (3.08) | -0.97 (3.24) | -1.48 (3.00) |
| White ethnicity, n (%)<br>[missing data for n=1,627] | 178,325 (96.8) | 19,803 (96.5) | 32,227 (97.7) | 49,562 (94.4) | 16,084 (98.0) |
| BMI [missing data for n=1879] | 26.8 (4.2) | 28.3 (5.1) | 27.7 (4.8) | 29.9 (5.2) | 27.7 (4.7) |
| Smoking, n (%) [missing data for n=1,965] |  |  |  |  |  |
| Never | 99,657 (54.1) | 10,286 (50.2) | 16,207 (49.2) | 23,017 (44.0) | 7,929 (48.4) |
| Previous | 68,417 (37.2) | 8,570 (41.8) | 13,358 (40.6) | 24,488 (46.8) | 7,052 (43.1) |
| Current | 16,034 (8.7) | 1,630 (8.0) | 3,380 (10.3) | 4,871 (9.3) | 1,393 (8.5) |
| Alcohol (weekly units)<br>[missing data for n=2,019] | 14.9 (17.9) | 13.3 (18.6) | 11.4 (16.7) | 15.4 (20.9) | 13.0 (17.3) |
| Alcohol group, n (%)<br>[missing data for n=2,019] |  |  |  |  |  |
| Low risk (<15 units/week) | 114,879 (62.4) | 13,840 (67.5) | 23,832 (72.4) | 32,962 (62.9) | 11,107 (67.8) |
| Increasing risk (14 – 35 units/week) | 58,710 (31.9) | 5,520 (26.9) | 7,591 (23.1) | 15,582 (29.8) | 4,407 (26.9) |
| High risk (>35 units/week) | 10,467 (5.7) | 1,138 (5.6) | 1,491 (4.5) | 3,840 (7.3) | 869 (5.3) |
| Physical Activity, n (%)<br>[missing data for n=4,282] |  |  |  |  |  |
| High | 15,307 (8.4) | 1,033 (5.1) | 1,309 (4.0) | 1,879 (3.6) | 767 (4.7) |
| Medium | 153,650 (84.1) | 16,455 (80.9) | 26,800 (81.8) | 41,491 (80.1) | 13,473 (82.8) |
| Low | 5,980 (3.3) | 1,032 (5.1) | 1,786 (5.5) | 3,026 (5.8) | 786 (4.8) |
| None | 7,875 (4.3) | 1,815 (8.9) | 2,855 (8.7) | 5,415 (10.5) | 1,238 (7.6) |
| Frailty<sup>1</sup>, n (%) [missing data for n=7,181] |  |  |  |  |  |
| Robust | 118,624 (65.4) | 9,569 (47.6) | 15,144 (46.8) | 21,707 (42.4) | 8,239 (51.1) |
| Pre-frail | 60,012 (33.1) | 9,131 (45.4) | 14,790 (45.7) | 24,777 (48.4) | 6,953 (43.2) |
| Frail | 2,688 (1.5) | 1,419 (7.1) | 2,422 (7.5) | 4,675 (9.1) | 923 (5.7) |
| Self-rated health, n (%)<br>[missing data for n=1,910] |  |  |  |  |  |
| Excellent | 40,566 (22.0) | 1,292 (6.3) | 2,111 (6.4) | 2,242 (4.3) | 1,263 (7.7) |
| Fair | 23,844 (12.9) | 6,608 (32.3) | 10,690 (32.5) | 19,743 (37.7) | 4,881 (29.9) |
| Good | 117,591 (63.8) | 10,786 (52.7) | 17,237 (52.4) | 24,716 (47.2) | 8,971 (54.9) |
| Poor | 2,302 (1.3) | 1,800 (8.8) | 2,854 (8.7) | 5,614 (10.7) | 1,228 (7.5) |

All data presented as mean (SD) unless otherwise stated. <sup>1</sup> Frailty according to the frailty phenotype.

#### UKHLS

### UKHLS: adults 18 – 36 years

**Table 8.** Sociodemographic characteristics of adults 18-36 years in UKHLS stratified by multimorbidity cluster.

|  | No multimorbidity | Pulmonary | Depression & Asthma | Cardiometabolic |
| --- | --- | --- | --- | --- |
| N (%) | 15,416 (95.7) | 270 (1.7) | 168 (1.0) | 251 (1.6) |
| Female, n (%) | 8,454 (54.8) | 176 (65.2) | 124 (73.8) | 175 (69.7) |
| Age | 27.3 (5.5) | 28.2 (5.4) | 27.9 (5.0) | 30.2 (4.8) |
| Number of LTCs<br>[from short list n=13] | 0.2 (0.4) | 2.3 (0.7) | 2.3 (0.5) | 2.3 (0.7) |
| White ethnicity, n (%) | 10,015 (68.3) | 226 (83.7) | 146 (86.9) | 213 (84.9) |
| BMI | 24.7 (4.8) | 27.1 (6.6) | 26.1 (6.7) | 28.1 (7.9) |
| Smoking *, n (%) |  |  |  |  |
| Never | 4,483 (46.8) | 64 (35.0) | 29 (24.8) | 65 (36.9) |
| Previous | 2,570 (26.9) | 52 (28.4) | 41 (35.0) | 38 (21.6) |
| Current | 2,520 (26.3) | 67 (36.6) | 47 (40.2) | 73 (41.5) |
| Alcohol *, n (%) |  |  |  |  |
| Never, or up to 2 times/year | 1,929 (24.4) | 41 (26.1) | 27 (29.0) | 46 (31.1) |
| Up to 1-2 times/month | 2,317 (29.3) | 51 (32.5) | 30 (32.3) | 50 (33.8) |
| 1-4 times/week | 3,221 (40.8) | 56 (35.7) | 29 (31.2) | 40 (27.0) |
| Daily or almost daily | 430 (5.5) | 9 (5.7) | 7 (7.5) | 12 (8.1) |
| Health-related quality of life |  |  |  |  |
| SF-12 Physical Component <sup>1</sup> | 54.0 (7.0) | 46.2 (11.7) | 48.0 (12.2) | 45.7 (12.8) |
| SF-12 Mental Component <sup>2</sup> | 50.4 (9.4) | 48.4 (10.2) | 37.1 (12.3) | 39.5 (13.1) |
| EQ-5D Index Score <sup>3</sup> | 0.88 (0.16) | 0.75 (0.25) | 0.72 (0.25) | 0.67 (0.29) |
| Self-rated health, n (%) |  |  |  |  |
| Excellent | 3,947 (25.7) | 25 (9.3) | 9 (5.4) | 8 (3.2) |
| Very good | 5,838 (37.9) | 56 (20.7) | 28 (16.7) | 47 (18.7) |
| Good | 4,078 (26.5) | 84 (31.1) | 56 (33.3) | 60 (23.9) |
| Fair | 1,212 (7.9) | 59 (21.9) | 45 (26.8) | 79 (31.5) |
| Poor | 313 (2.0) | 46 (17.0) | 30 (17.9) | 57 (22.7) |

\*Smoking and alcohol were not collected at baseline, data presented here from second wave of data collection. <sup>1</sup> Score of <50 recommended as cut off for determining a 'physical condition'. <sup>2</sup> Score < 42 may be indicative of clinical depression. <sup>3</sup> EQ-5D index score mapped from SF-12 using.

### UKHLS: adults 37 – 54 years

**Table 9.** Sociodemographic characteristics of adults 37 - 54 years in UKHLS stratified by multimorbidity clusters.

|  | No multimorbidity | Pulmonary | Arthritis+ | Diabetes & Hypertension | Cardiovascular | Cancer, Thyroid disease & Depression |
| --- | --- | --- | --- | --- | --- | --- |
| N (%) | 14,542 (86.9) | 805 (4.8) | 466 (2.8) | 210 (1.3) | 218 (1.3) | 485 (2.9) |
| Female, n (%) | 7,758 (53.4) | 553 (68.7) | 313 (67.2) | 109 (51.9) | 98 (45.0) | 315 (65.0) |
| Age | 44.8 (5.0) | 45.8 (5.2) | 47.8 (4.8) | 47.8 (5.0) | 47.8 (4.8) | 46.5 (5.1) |
| Number of LTCs<br>[from short list n=13] | 2.9 (3.7) | 2.6 (0.9) | 2.4 (0.7) | 2.4 (0.5) | 3.2 (1.4) | 2.3 (0.6) |
| White ethnicity, n (%) | 11,054 (78.9) | 693 (86.2) | 388 (83.3) | 108 (51.4) | 174 (79.8) | 414 (85.4) |
| BMI | 26.5 (4.9) | 28.4 (6.4) | 28.9 (6.7) | 31.3 (6.5) | 29.8 (7.5) | 28.8 (6.6) |
| Smoking*, n (%) |  |  |  |  |  |  |
| Never | 5,027 (47.4) | 220 (35.4) | 123 (34.0) | 91 (59.9) | 46 (28.1) | 141 (36.7) |
| Previous | 3,266 (30.8) | 204 (32.9) | 124 (34.3) | 33 (21.7) | 47 (28.7) | 120 (31.3) |
| Current | 2,321 (21.9) | 197 (31.7) | 115 (31.8) | 28 (18.4) | 71 (43.3) | 123 (32.0) |
| Alcohol*, n (%) |  |  |  |  |  |  |
| Never up to 2 times/year | 1,786 (19.6) | 149 (28.6) | 95 (30.1) | 47 (42.7) | 43 (33.1) | 99 (30.1) |
| Up to 1-2 times/month | 2,057 (22.5) | 145 (27.8) | 79 (25.0) | 29 (26.4) | 34 (26.2) | 75 (22.8) |
| 1-4 times/week | 4,116 (45.1) | 162 (31.1) | 97 (30.7) | 28 (25.5) | 42 (32.3) | 115 (35.0) |
| Daily or almost daily | 1,172 (12.8) | 65 (12.5) | 45 (14.2) | 6 (5.5) | 11 (8.5) | 40 (12.1) |
| Health-related quality of life |  |  |  |  |  |  |
| SF-12 Physical Component <sup>1</sup> | 52.4 (8.8) | 42.3 (13.5) | 37 (13.4) | 42.2 (12.3) | 33.9 (13.1) | 44.0 (13.0) |
| SF-12 Mental Component <sup>2</sup> | 50.4 (9.5) | 43.7 (13.5) | 43.3 (13.6) | 46.9 (11.4) | 41.8 (13.3) | 41.4 (13.1) |
| EQ-5D Index Score <sup>3</sup> | 0.85 (0.19) | 0.66 (0.31) | 0.57 (0.32) | 0.71 (0.26) | 0.53 (0.34) | 0.67 (0.31) |
| Self-rated health, n (%) |  |  |  |  |  |  |
| Excellent | 3,063 (21.1) | 42 (5.2) | 16 (3.4) | 5 (2.4) | 3 (1.4) | 22 (4.5) |
| Very good | 5,196 (35.8) | 125 (15.5) | 52 (11.2) | 18 (8.6) | 13 (6.0) | 69 (14.2) |
| Good | 4,203 (28.9) | 207 (25.7) | 95 (20.4) | 66 (31.4) | 34 (15.6) | 130 (26.8) |
| Fair | 1,595 (11.0) | 229 (28.5) | 163 (35.0) | 65 (31.0) | 61 (28.0) | 148 (30.5) |
| Poor | 464 (3.2) | 202 (25.1) | 140 (30.0) | 56 (26.7) | 107 (49.1) | 116 (23.9) |

\*Smoking and alcohol data were not collected at baseline, data presented here from second wave of data collection.
<sup>1</sup> Score of <50 recommended as cut off for determining a 'physical condition'. <sup>2</sup> Score < 42 may be indicative of clinical depression. <sup>3</sup> EQ-5D index score mapped from SF-12.

### UKHLS: adults 55 - 73 years

**Table 10.** Sociodemographic characteristics of adults 55 - 73 years in UKHLS stratified by multimorbidity clusters.

|  | No multimorbidity | Pulmonary | Hypertension+ | Cardiovascular | Cancer, Thyroid disease & Depression |
| --- | --- | --- | --- | --- | --- |
| N (%) | 8,191 (66.6) | 730 (5.9) | 2,162 (17.6) | 584 (4.7) | 641 (5.2) |
| Female, n (%) | 4,218 (51.5) | 478 (65.5) | 1,198 (55.4) | 226 (38.7) | 436 (68.0) |
| Age | 62.7 (5.2) | 63.4 (5.1) | 64.6 (5.3) | 65.4 (5.3) | 63.1 (5.1) |
| Number of LTCs<br>[from short list n=13] | 0.5 (0.5) | 2.84 (1.0) | 2.6 (0.9) | 3.5 (1.5) | 2.3 (0.6) |
| White ethnicity, n (%) | 7,120 (90.5) | 684 (93.7) | 1,913 (88.6) | 529 (90.7) | 606 (94.5) |
| BMI | 26.4 (4.4) | 27.9 (5.5) | 28.8 (5.3) | 28.9 (5.9) | 27.4 (5.4) |
| Smoking, n (%) |  |  |  |  |  |
| Never | 2,547 (39.9) | 189 (30.9) | 646 (36.3) | 120 (26.8) | 188 (36.2) |
| Previous | 2,816 (44.1) | 299 (48.9) | 856 (48.1) | 232 (51.8) | 224 (43.1) |
| Current | 1,016 (15.9) | 124 (20.3) | 279 (15.7) | 96 (21.4) | 108 (20.8) |
| Alcohol frequency, n (%) |  |  |  |  |  |
| Never, or up to 2 times/year | 1,127 (19.8) | 169 (30.8) | 459 (29.1) | 153 (40.7) | 132 (28.6) |
| Up to 1-2 times/month | 1,029 (18.0) | 115 (21.0) | 299 (19.0) | 64 (17.0) | 110 (23.8) |
| 1-4 times/week | 2,454 (43.0) | 180 (32.9) | 519 (32.9) | 108 (28.7) | 151 (32.7) |
| Daily or almost daily | 1,095 (19.2) | 84 (15.3) | 300 (19.0) | 51 (13.6) | 69 (14.9) |
| Health-related quality of life |  |  |  |  |  |
| SF-12 Physical Component <sup>1</sup> | 49.7 (10.3) | 38.4 (14.0) | 38.7 (13.5) | 30.7 (13.1) | 39.3 (13.7) |
| SF-12 Mental Component <sup>2</sup> | 52.9 (8.7) | 48.9 (12.4) | 50.4 (11.3) | 48.2 (12.6) | 47.9 (12.6) |
| EQ-5D Index Score <sup>3</sup> | 0.84 (0.20) | 0.64 (0.31) | 0.68 (0.29) | 0.55 (0.33) | 0.65 (0.30) |
| Self-rated health, n (%) |  |  |  |  |  |
| Excellent | 1,441 (17.7) | 25 (3.4) | 68 (3.2) | 2 (0.3) | 27 (4.2) |
| Very good | 2,771 (33.9) | 115 (15.8) | 310 (14.4) | 41 (7.0) | 105 (16.4) |
| Good | 2,378 (29.1) | 193 (26.5) | 667 (30.9) | 84 (14.4) | 185 (28.9) |
| Fair | 1,184 (14.5) | 212 (29.1) | 677 (31.3) | 187 (32.0) | 195 (30.4) |
| Poor | 390 (4.8) | 184 (25.2) | 439 (20.3) | 270 (46.2) | 129 (20.1) |

\*Smoking and alcohol data were not collected at baseline, data presented here from second wave of data collection. <sup>1</sup> Score of <50 recommended as cut off for determining a 'physical condition'. <sup>2</sup> Score < 42 may be indicative of clinical depression. <sup>3</sup> EQ-5D index score mapped from SF-12.

### UKHLS: adults 74+ years

**Table 11.** Sociodemographic characteristics of adults 74+ years in UKHLS stratified by multimorbidity clusters.

|  | No multimorbidity | Pulmonary | Arthritis+ | Cardiometabolic |
| --- | --- | --- | --- | --- |
| N (%) | 2,161 (53.4) | 380 (9.4) | 959 (23.7) | 547 (13.5) |
| Female, n (%) | 1,131 (52.3) | 232 (61.1) | 633 (66.0) | 230 (42.1) |
| Age | 80.1 (5.0) | 79.8 (4.5) | 80.2 (4.8) | 79.7 (4.8) |
| Number of LTCs<br>[from short list n=13] | 0.6 (0.5) | 3.4 (1.3) | 2.8 (1.0) | 2.4 (0.7) |
| White ethnicity, n (%) | 1,900 (94.0) | 356 (93.7) | 901 (94) | 510 (93.2) |
| BMI | 25.2 (4.1) | 26.6 (5.2) | 26.9 (5.0) | 26.2 (4.6) |
| Smoking*, n (%) |  |  |  |  |
| Never | 653 (43.1) | 94 (32.4) | 304 (42.2) | 155 (37.4) |
| Previous | 752 (49.6) | 169 (58.3) | 371 (51.5) | 237 (57.3) |
| Current | 110 (7.3) | 27 (9.3) | 46 (6.4) | 22 (5.3) |
| Alcohol*, n (%) |  |  |  |  |
| Never up to 2 times/year | 388 (31.0) | 98 (41.7) | 234 (39.4) | 119 (35.4) |
| Up to 1-2 times/month | 242 (19.3) | 50 (21.3) | 114 (19.2) | 75 (22.3) |
| 1-4 times/week | 394 (31.5) | 49 (20.9) | 150 (25.3) | 87 (25.9) |
| Daily or almost daily | 228 (18.2) | 38 (16.2) | 96 (16.2) | 55 (16.4) |
| Health-related quality of life |  |  |  |  |
| SF-12 Physical Component <sup>1</sup> | 43.1 (12.4) | 30.4 (12.5) | 31.1 (12.4) | 37.1 (12.8) |
| SF-12 Mental Component <sup>2</sup> | 53.1 (9.4) | 50.0 (11.5) | 51.2 (11.5) | 51.7 (10.4) |
| EQ-5D Index Score <sup>3</sup> | 0.78 (0.24) | 0.57 (0.32) | 0.60 (0.29) | 0.70 (0.26) |
| Self-rated health, n (%) |  |  |  |  |
| Excellent | 261 (12.2) | 6 (1.6) | 31 (3.2) | 14 (2.6) |
| Very good | 579 (26.9) | 41 (10.8) | 140 (14.6) | 80 (14.7) |
| Good | 636 (29.6) | 63 (16.6) | 217 (22.7) | 150 (27.5) |
| Fair | 438 (20.4) | 129 (34.0) | 327 (34.1) | 189 (34.7) |
| Poor | 235 (10.9) | 141 (37.1) | 243 (25.4) | 112 (20.6) |

\*Smoking and alcohol data were not collected at baseline, data presented here from second wave of data collection. <sup>1</sup> Score of <50 recommended as cut off for determining a 'physical condition'. <sup>2</sup> Score < 42 may be indicative of clinical depression. <sup>3</sup> EQ-5D index score mapped from SF-12.

## Discussion

Using LCA we identified latent classes of LTCs across three databanks and four age groups. Clusters identified in different age groups in all databanks center around specific systems in the body. For example, in all age groups and databanks LCA identified clusters centring around pulmonary conditions. Other common clusters combine cardiometabolic diseases, mental health or neurological conditions, or painful conditions. Although the exact combination of LTCs in clusters necessarily differ depending on the data used for LCA, previous studies applying LCA to different cohorts or databanks have likewise consistently identified clusters combining conditions around different systems including among others, cardiometabolic clusters, pain-related clusters, respiratory clusters, and neurological/mental health clusters [13–17]. In addition to clusters which group disorders affecting specific systems, discordant clusters are clusters grouping disorders with different aetiologies and affected systems which may have a more complex interpretation. Despite the differences in details on how many conditions were considered and how the conditions were entered into the databanks and cohorts between previous studies, as well as between the different databanks in this study, similar types of clusters tend to emerge across cohorts/databanks. These similarities in clusters suggest that there might be important associations and potentially connected aetiologies between certain LTCs, leading them to cluster together more frequently.

## Data Availability

Access to data from Biobank can be requested via the UK Biobank Access Management System https://www.ukbiobank.ac.uk/enable-your-research/apply-for-access
Access to SAIL data can be requested via the independent Information Governance Review Panel https://saildatabank.com/data/apply-to-work-with-the-data/
Access to UKHLS data can be requested via UK Data Service https://www.understandingsociety.ac.uk/documentation/access-data

https://saildatabank.com/data/apply-to-work-with-the-data/

https://www.understandingsociety.ac.uk/documentation/access-data

